# The impact of supervised beego, a traditional Chinese water-only fasting, on thrombosis and hemostasis

**DOI:** 10.1101/2020.07.31.20166215

**Authors:** Yixuan Fang, Yue Gu, Chen Zhao, Yaqi Lv, Jiawei Qian, Lingjiang Zhu, Na Yuan, Suping Zhang, Li Wang, Mengli Li, Qing Zhang, Li Xu, Wen Wei, Lei Li, Li Ji, Xueqin Gao, Jingyi Zhang, Yueping Shen, Zixing Chen, Guanghui Wang, Kesheng Dai, Jianrong Wang

**Affiliations:** Hematology Center of Cyrus Tang Medical Institute, Key Laboratory of Thrombosis and Hemostasis of Ministry of Health, National Clinical Research Center for Hematologic Diseases, Collaborative Innovation Center of Hematology, Jiangsu Institute of Hematology, Institute of Blood and Marrow Transplantation, Department of Hematology, The First Affiliated Hospital of Soochow University School of Medicine, Suzhou 215123, China; Soyo Center, Soochow University, Suzhou 215123, China; State Key Laboratory of Radiation Medicine and Radioprotection, Soochow University, Suzhou 215123, China; Department of Community Nursing, Soochow University School of Nursing, Suzhou 215006, China; Department of Kinesiology, Soochow University School of Physical Education and Sport Sciences, Suzhou 215021, China; Department of Epidemiology and Biostatistics, Soochow University School of Public Health, Suzhou 215123, China; Department of Pharmacology, Soochow University School of Pharmacy, Suzhou 215123, China

## Abstract

Beego is a traditional Chinese complete water-only fasting practice initially developed for spiritual purposes, later extending to physical fitness purposes. Beego notably includes a psychological induction component that includes meditation and abdominal breathing, light body exercise, and ends with a specific gradual refeeding program before returning to a normal diet. Beego has regained its popularity in recent decades in China as a strategy for helping people in subhealthy conditions or with metabolic syndrome, but we are unaware of any studies examining the biological effects of this practice. To address this, we here performed a longitudinal study of beego comprising fasting (7 and 14 day cohorts) and a 7-day programmed refeeding phase. In addition to detecting improvements in cardiovascular physiology and selective reduction of blood pressure in hypertensive subjects, we observed that beego decreased blood triacylglycerol (TG) selectively in TG-high subjects and increased cholesterol in all subjects during fasting; however, the cholesterol levels were normalized after completion of the refeeding program. Strikingly, beego reduced platelet formation, activation, aggregation, and degranulation, resulting in an alleviated thrombosis risk, yet maintained hemostasis by sustaining levels of coagulation factors and other hemostatic proteins. Mechanistically, we speculate that downregulation of G6B and MYL9 may influence the observed beego-mediated reduction in platelets. Fundamentally, our study supports that supervised beego reduces thrombosis risk without compromising hemostasis capacity. Moreover, our results support that beego under medical supervision can be implemented as noninvasive intervention for reducing thrombosis risk, and suggest several lines of intriguing inquiry for future studies about this fasting practice (http://www.chictr.org.cn/index.aspx, number,ChiCTR1900027451).

## Introduction

Beego, a phonetic articulation in Chinese meaning no eating foods for a period, was originally practiced by Taoists for spiritual purposes in ancient China. It started from the Qin Dynasty (from 221 B.C. to 207 B.C.) and became a popular practice in the Western Han Dynasty (206 B.C. to 24 A.D.), extending to the purposes for both physical and mental fitness. Like Gongfu or martial arts, beego is a part of traditional Chinese culture in fitness. But unlike Gongfu that requires a certain physical talent, beego can be practiced by ordinary people. It consists of two phases, starting with a complete water-only fasting phase for days or weeks, followed by a gradual refeeding phase for the duration of no less than half of the fasting length till normal diet. Psychological induction or mind regulation, including but not limited to meditation, is used to mitigate the feeling of hunger, in particular in the first days of fasting. During meditation, an abdominal breathing method is used to inhale sufficient oxygen and exhale more carbon dioxide^1,2^. In addition, light body exercise is incorporated in beego program to facilitate this practice. In recent two decades with rapid increase of the population in overnutritional condition or with metabolic syndrome, there is also a rapid growing volume of Chinese people participating in beego.

Beego in China has traditionally relied on experience being handed down by word of mouth and features a psychological guidance, yet we are unaware of studies examining the practice; in contrast fasting for therapeutic purposes in the West has been intensively studied since the early 1900s, including modalities like prolonged or acute starvation^3-6^, water-only fasting^7-10^ and various less stringent or incomplete fasting formulas^11^. The incomplete fasting formulas appear to be more favored in Western countries, and include calorie restriction, intermittent fasting, fasting-mimicking diets and periodic short-term water-only fasting with minimal calories or supplements. Contrary to the difficulties in studying complete fasting biology that precludes utilization of animals (as some animals cannot survive complete fasting for days), studies on incomplete fasting primarily using model rodents has revealed health-promoting physiologic responses, and have uncovered underlying mechanisms that can explain these responses. These include but not limited to ketogenesis^12-14^, hormone modulation^15-17^, reduced inflammation^18^, immunological memory^19^, promotes regeneration and reduces autoimmunity and multiple sclerosis symptoms^20^, and fasting confers protection in CNS autoimmunity by altering the gut microbiota^21^. Fasting also impacts immune cell dynamics and mucosal immune responses^22^, and increases stress resistance^23^, lipolysis^24^, and autophagy^25,26^. Caloric restriction delays disease onset and mortality in rhesus monkeys^27,28^.

More encouragingly, clinical investigations in Western countries indicate that fasting reduces obesity^3^ and hypertension^7,8^, improves oxidative stress^29,30^, cardiovascular disease^31-33^, cancer^34-38^, rheumatoid arthritis^39^, metabolic syndrome^40,41^, osteoarthritis^42^, fibromyalgia^43^, chronic pain^44^, memory^45^, circadian clock^46^ and multiple aspects of life quality^11,47-50^.

However, the safety and the biological effects of beego, the Chinese traditional fasting practice, remain completely undocumented in scientific format. This study aimed to explore the impact of supervised beego on thrombosis and hemostasis.

## Results

### 1. Supervised beego alleviates thrombosis risk after water-only fasting and refeeding

The protocol for beego is detailed in the methods. Comparisons were made between the baseline value tested on the beginning on day 1 (wFD1) and the indicated testing days (Figure 1a; Supplementary Figure S1a). Blood lipids were measured over the course of beego program at 4 or 5 time points, respectively, for the 7-day fasting or 14-day fasting programs. For subjects who have normal triacylglycerol (TG) level before beego, TG values were maintained within a normal range throughout the entire fasting and refeeding periods; although there were variations evident on fasting days 4 and 7, the levels normalized to the baseline upon completion of the 7-day refeeding (Figure 1b, left). In contrast, for TG high subjects, the TG level decreased, reaching values within a normal range during fasting; this decrease was sustained in the normal range on refeeding day 7 (Figure 1b, right). Note that these patterns for normal and high TG subjects were also consistent for the 14-day fasting program (Supplementary Figure S1b).

**Figure 1.**
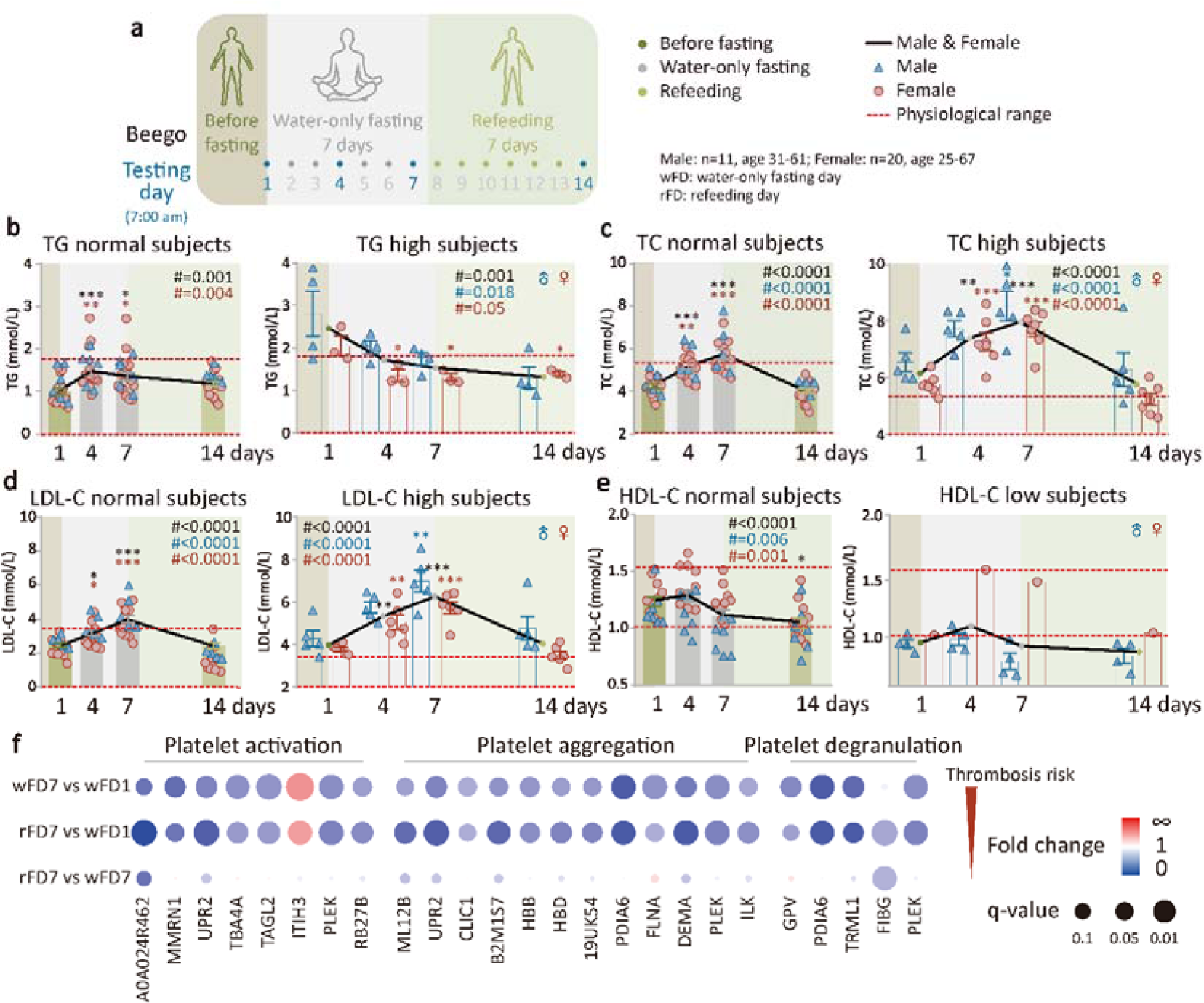
Supervised beego alleviates thrombosis risk after water-only fasting and refeeding. **a**. The Beego protocol used in the present study comprised seven complete water-only fasting days, associated with psychological induction and breath training, followed by seven refeeding days (rFD). Testing was performed at 7:00-9:00 am of the following days: wFD 1, 4, 7, and rFD 7 as indicated. **b**. blood triacylglycerol (TG) test in TG normal (male=6, female=16) and high (male=4, female=3) subjects. **c**. Blood total cholesterol (TC) test in TC normal (male=5, female=12) and TC high (male=5, female=7) subjects. **d**. Low-density lipoprotein cholesterol (LDL-C) test in LDL-C normal (male=5, female=13) and high (male=5, female=6) subjects. **e**. HDL-C in the normal (male=7, female=10) and low (male=4, female=1) subjects. **f**. Bubble diagram of plasma proteomics analysis (label-free data-independent acquisition) for ten subjects for each group (5 for male and female each, aged 30-60). GO enrichment analysis for three categories of proteins responsible for platelet-related thrombosis risk, with comparison between the testing days as indicated. Fold change is shown by the color and q-value for statistical significance is represented by the size of the dots. Data are means ± SEM. \**P* < 0.05; \*\**P* < 0.01; \*\*\**P* < 0.001. *significant compared with the wFD1 immediately before the start of fasting.

For subjects with normal total cholesterol (TC) levels before beego, the TC level remained within a normal range, despite an increase during the 7-day fasting period; the TC level decreased to a level close to the baseline after 7-day refeeding (Figure 1c, left). However, for high TC subjects, although TC increased significantly on fasting day 4 and 7, it decreased to the baseline after 7-day refeeding (Figure 1c, right). Furthermore, 14-day fasting following 7-day refeeding resulted in a significant reduction in TC levels for all subjects, including the TC normal and high subjects, with final values close to the midline of the normal TC range (Supplementary Figure S1c).

For low-density lipoprotein cholesterol (LDL-C) normal subjects, LDL-C increased toward the upper range of normal values during fasting, but returned to the baseline after 7-day refeeding (Figure 1d, left). For high LDL-C subjects, LDL-C significantly increased along with fasting but returned to the baseline after 7-day refeeding (Figure 1d, right). Similar to the above metabolic factors, high-density lipoprotein cholesterol (HDL-C), a positive marker for health, was reduced to within normal values in the fasting and refeeding periods for the HDL-C normal subjects (Figure 1e, left); but in the low HDL-C subjects, beego did not cause significant change in HDL-C level (Figure 1e, right). Interestingly, in the first seven days of fasting period, TG reduction (good for health) and total cholesterol increase (previously regarded as largely bad for health) displayed opposing trends in both the metabolically normal and abnormal subjects. After 7-day fasting, both the TC and TG levels were progressively reduced, with the exception that LDL-C that was not reduced until completion of the 7-day fasting. Furthermore, 14-day fasting followed by 7-day refeeding also showed a better pattern in the dynamic changes of blood lipids (Supplementary Figure S1c,d,e). Together, these results suggest that while beego apparently decreases TG-trigged thrombosis risk factors, it may also transiently increase the cholesterol-triggered thrombosis risk during fasting period. This may reflect the glucogenesis-to-ketogenesis (G-K) switch in the body’s energy supply^12^. However, it should be noted that we found this transient increase in TC during fasting is reversible by refeeding, and 14-day fasting following 7-day refeeding results in further reduction in TC and LDL-C levels.

Apart from blood lipids, abnormally elevated platelet activation and aggregation are major risks for thrombosis^51,52^. To determine if beego enhances any platelet contribution to thrombosis, we performed a plasma proteomics analysis of the subjects before fasting (wFD1), after 7-day fasting (wFD7), and after 7-day refeeding (rFD7). Fundamentally, this analysis revealed that the expression of proteins involved in platelet activation, platelet aggregation, and platelet degranulation was significantly decreased at the end of 7-day fasting as compared to the baseline before fasting. Moreover, the data for the 7 days refeeding timepoint supported that this reduction was sustained (Figure 1f). Since platelet degranulation and aggregation are known to contribute to the process of platelet activation^52^, these results together suggest a reduced risk of platelet-triggered thrombosis.

### 2. Supervised beego maintains hemostasis capacity

Administration of anti-thrombosis drugs such as aspirin is known to cause stomach bleeding^53^. It is thus necessary to examine if beego reduces thrombosis risk at a cost of compromising hemostasis capacity. Hemostasis is a tightly regulated process of coagulation that prevents and stops bleeding while maintaining normal blood flow in the vessels. The coagulation cascade comprising two primary pathways (intrinsic and extrinsic pathways). The intrinsic pathway involves factors XII, XI, IX, VIII, and V, while the extrinsic pathway involves factors VII, X, and prothrombin. Both pathways eventually meet and together accomplish clot production. This common, final phase involves factors I, II, V, and X. In our study, plasma levels of proteins involved in coagulation cascades were tested by liquid chromatography-tandem mass spectrometry at three time points (wFD1, wFD7 and rFD7) from 5 male and 5 female subjects. We found that the factors in both of the intrinsic and the extrinsic pathways maintained similar levels in the fasting and refeeding beego phases (Figure 2a). Further, a plasma proteomics study also indicated a lack of any obvious changes for 16 known hemostasis-related proteins in fasting and refeeding as compared with the baseline before fasting (Figure 2b). It thus appears that beego does not obviously modulate pro-coagulants or anti-coagulants.

**Figure 2.**
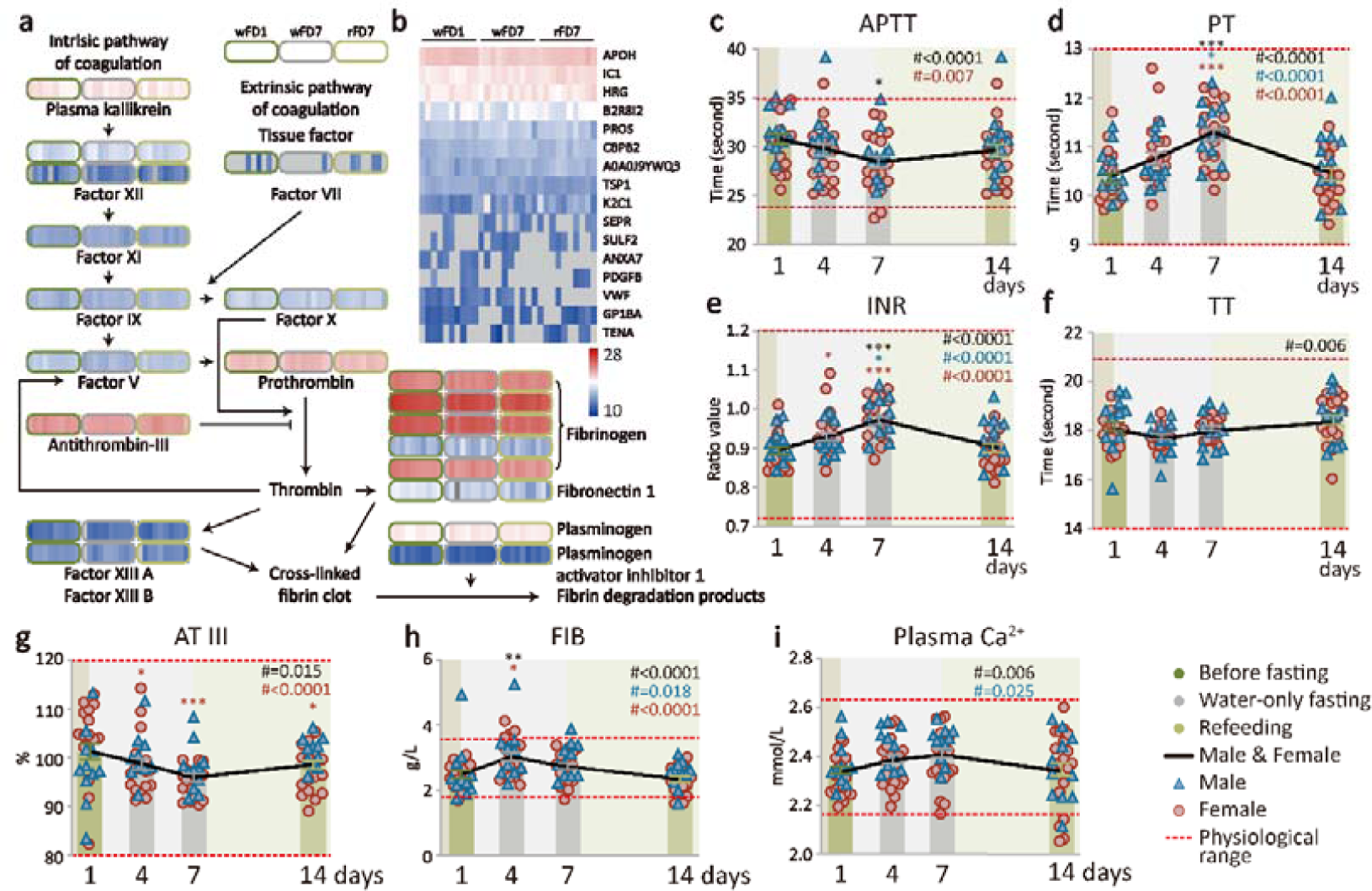
Supervised beego maintains hemostasis capacity. **a**. Heatmap with coagulation cascades data obtained via label-free data-independent acquisition. Plasma levels of proteins involved in coagulation cascades were tested by liquid chromatography-tandem mass spectrometry at three time points (wFD1, wFD7, and rFD7) from five males and five females for each group. **b**. Heatmap of hemostasis related proteins. The levels of proteins were tested at three time points (wFD1, wFD7, and rFD7) from 5 males and 5 females. The grey positions represent proteins with abundance too low to be reliably assessed as enriched. **c-f**. Coagulation tests. Clotting dynamics were measured at four time points (wFD1, wFD4, wFD7, and rFD7). The tests include activated partial thromboplastin time (APTT, male=11, female=17), prothrombin time (PT, male=11, female=17), internal normalized ratio (INR, male=11, female=17), thrombin time (TT, male=11, female=17). **g-i**. Plasma levels of hemostatic biomarkers. The levels of known hemostasis-related proteins were measured at four time points as indicated. The tests included antithrombin III (AT III, male=11, female=17); fibrinogen (FIB, male=11, female=17); Ca^2+^ in plasma, male=11, female=20. Data are means ± SEM. \**P* < 0.05; \*\**P* < 0.01; \*\*\**P* < 0.001. *significant compared with the wFD1 (7:00 am), immediately before the start of fasting.

Seeking further support to bolster the proteomics result indicating that beego may reduce thrombosis risk while maintaining hemostasis capacity, we performed laboratory tests to evaluate coagulation function. Activated partial thromboplastin time (APTT) measures the time it takes for a clot to form, and examines the integrity of the intrinsic system and common factors. The APTT of beego subjects progressively decreased within the normal range during fasting; further, the APTT was maintained at around 30 seconds upon completion of refeeding, a value in the middle of the normal range (Figure 2c). Prothrombin time (PT) measures how quickly blood clots by specifically evaluating the presence of factors VII, V, and X, prothrombin, and fibrinogen, thus reflecting the integrity of the extrinsic system as well as factors common to both systems. PT increased during fasting within normal range and normalized to the values between 10.5 and 11.3 seconds after refeeding for the beego subjects, well within the normal PT range (Figure 2d). We also noted that beego subject values for the international normalized ratio (INR)—which is expressed as a ratio of PT—were also increased during fasting and returned to within a normal range (0.9 to 0.98) during refeeding (Figure 2e). Thrombin time (TT) test measures fibrin formation, the final reaction of the clotting cascade. Beego subject TT values remained between 18 and 18.5 seconds, well within the normal range (Figure 2f). Note that these four hemostasis parameters were all well normalized to the baseline after beego.

The beego subjects exhibited decreased antithrombin III (AT III) activity during fasting but it was normalized during refeeding; nevertheless, AT value of the beego subjects remained around 100 seconds, close to the middle value of the normal range throughout the beego program (Figure 2g). Fibrinogen (FIB) is one of 13 coagulation factors responsible for normal blood clotting. During the entire fasting and refeeding period, the FIB values were maintained around 3 mmol/L, well within the normal range (Figure 2h). Calcium ions in plasma are required for activation of blood clotting pathways. The concentration of calcium ions in plasma of beego subjects was sustained between 2.3 and 2.4 mmol/L, well within normal range (Figure 2i). Note that beego comprising 14-day fasting and 7-day refeeding displayed a similar pattern for dynamic changes in hemostasis parameters (Supplementary Figure S2). These results together suggest that the activity and amount of thrombin and antithrombin III remain unchanged upon completion of the beego program.

### 3. Supervised beego improves cardiovascular physiology

Intact blood vessels are instrumental to limiting thrombosis and moderating the capacity of platelets to secure hemostasis^54-56^. Therefore, the physiology of vessels is fundamental for supporting hemostasis. Peripheral resistance measures vascular compliance, reflecting friction between the blood and the walls of the blood vessels: both narrower blood vessels and relatively more viscous blood increase peripheral resistance. We observed that the peripheral resistance of beego subjects was maintained in the middle of the normal range during fasting, yet decreased during refeeding, while remaining within the normal range (Figure 3a). The reflection coefficient data showed a slow, progressive decrease during fasting, with a further decrease evident after refeeding (Figure 3b), results suggesting improved vascular function. The augmentation index (AIx) is an indicator of peripheral arterial stiffness. This increased within normal range during fasting, but decreased close to the baseline value after refeeding (Figure 3c). Pulse wave velocity (PWV) is a measure of aortic arterial stiffness that is a potent predictor of cardiovascular risk; we observed no change in PWV for beego subjects during fasting or refeeding (Figure 3d). These data together suggest that vascular-dysfunction-related hypertension risk was reduced in beego subjects by day 7 of refeeding.

**Figure 3.**
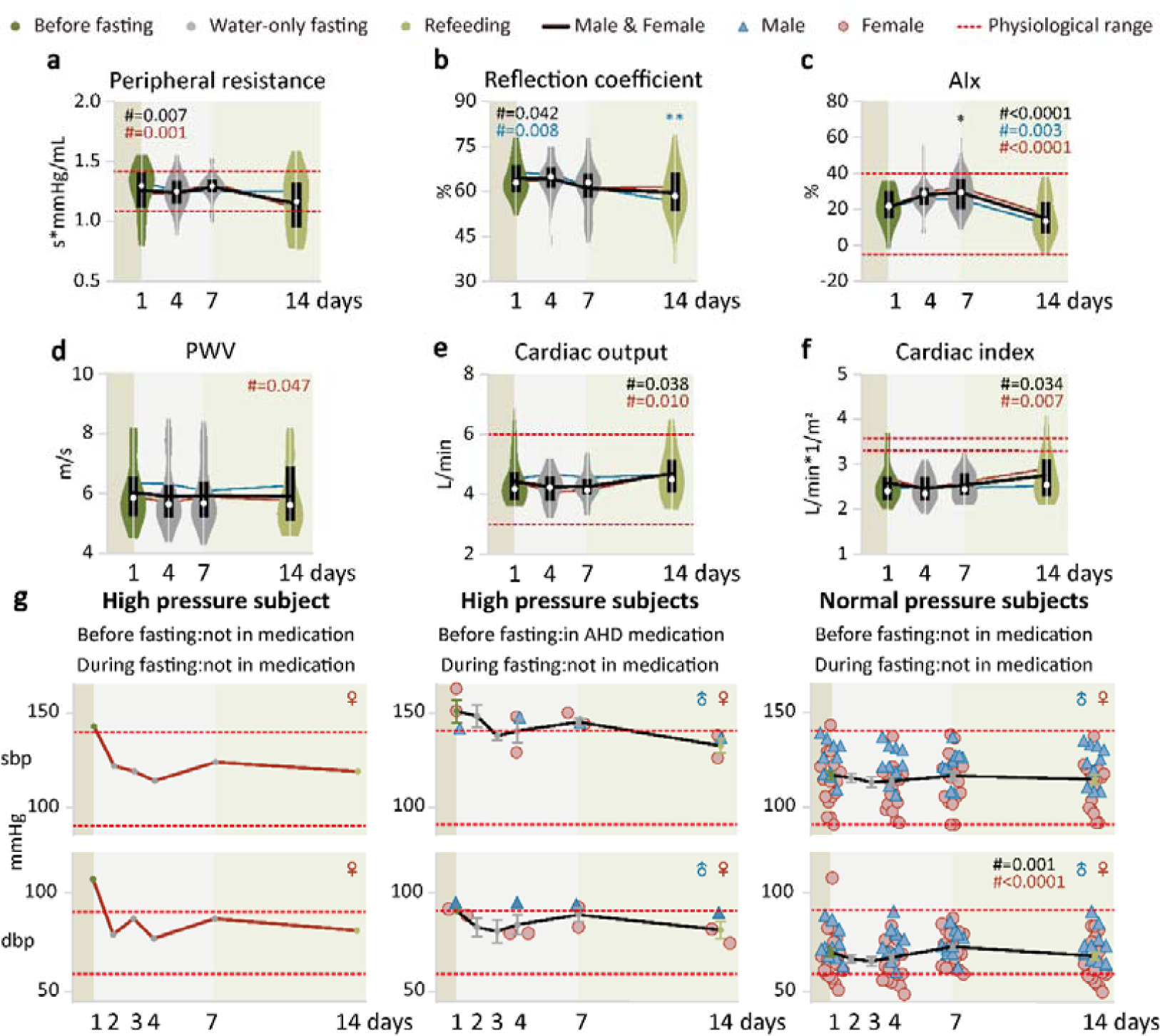
Supervised beego improves cardiovascular physiology. **a-d**. Violin plots of vascular health measured with Mobil-O-Graph PWA monitor. Measurements include peripheral resistance (male=10, female=18), reflection coefficient (male=10, female=18), augmentation index (Alx, male=10, female=18), and pulse wave velocity (PWV, male=10, female=18). **e-f**. Violin plots of cardiac health measured with Mobil-O-Graph PWA. Measurements include cardiac output (male=10, female=18) and cardiac index (male=10, female=18). **g**. Blood pressure measurement in hypertension or normal blood pressure groups. Left panel, hypertensive subject not on medication before or during fasting or refeeding (female=1); middle panel, hypertensive subjects on antihypertension medication before fasting but not taking the medication during fasting or refeeding (male=1, female=2); right panel, normal blood pressure subjects not on medication (male=10, female=18). sbp=systolic blood pressure, dbp=diastolic blood pressure. Data are means ± SEM. \**P* < 0.05; \*\**P* < 0.01; \*\*\**P* < 0.001. *significant compared with the wFD1 (7:00 am) immediately before the start of fasting.

In cardiovascular physiology, cardiac output describes the volume of blood being pumped the left and right ventricle of the heart per unit time. Cardiac output remained unchanged during fasting but slightly increased after refeeding (Figure 3e). Cardiac index is defined as cardiac output/body surface area, and monitoring of this parameter allows comparisons of cardiac function between individuals. After completion of beego, the cardiac index maintained unchanged (Figure 3f). We noted improvement of vascular function as indicated by decreases in systolic (SBP) and diastolic blood pressure (DBP) in the hypertensive subjects (none of whom were taking antihypertensioin drugs during beego) (Figure 3g, left panel). Importantly, all of the hypertensive subjects who had taken antihypertension medication before fasting were able to maintain normal blood pressure without medication during beego; moreover the blood pressure values achieved with beego (without medication) appeared to be lower than the baseline values for these medicated subjects before beego (Figure 3g, middle panel). For the normal blood pressure subjects, their blood pressure maintained quite stable and within the normal range before and after beego (Figure 3g, right panel). Beego with 14-day fasting and 7-day refeeding displayed similar patterns for these cardiovascular parameters (Supplementary Figure S3). Thus, beego can reduce blood pressure in hypertensive individuals and promotes cardiovascular health.

### 4. Supervised beego reduces platelet counts but retains platelet function

Platelets play an important role in the initiation of blood clotting^57,58^. To examine if production of platelets in the body responds to beego, we measured peripheral platelet count at various beego time points. Notably, for subjects with normal platelet counts, fasting caused a progressive decrease in its count to a low value within the normal range (Figure 4a, left). Low platelet count appears to be more frequently seen in females: all five of the low platelet subjects in the study were female. Surprisingly, fasting did not reduce platelet count in the low platelet count subjects; instead, it led to an increasing tendency for the average platelet count, and this increase in average platelet number was maintained after 7 days of refeeding (Figure 4a, right). This finding underscores that the body’s response to beego is highly suitable: it induces a reduction in the platelet pool if there are sufficient platelets or promotes the maintenance (or even enhancement) of platelet generation if there are insufficient platelets.

**Figure 4.**
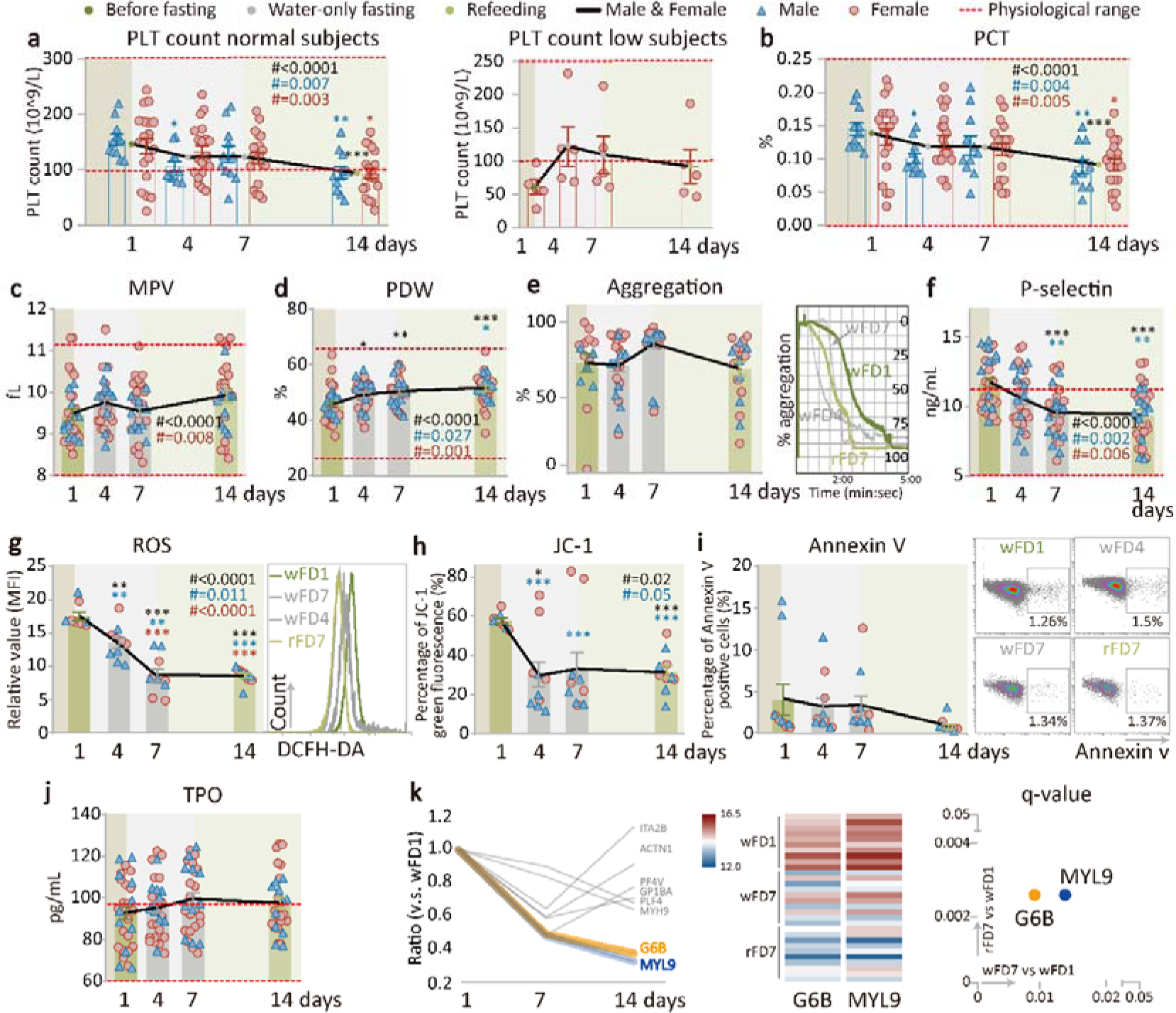
Supervised beego reduces platelet counts but retains platelet function. **a**. Platelet (PLT) count in PLT normal (male=11, female=20) or low groups (female=5). **b-d**. Platelet hematocrit (PCT, male=11, female=20), mean platelet volume (MPV, male=11, female=20), platelet distribution width (PDW, male=11, female=20). e. Platelet aggregation test. Left panel, platelet aggregation after stimulation was tested on the indicated days; right panel, representative aggregation tracing for thrombin-induced platelet activation was monitored using platelet aggregometry (male=11, female=20). f. ELISA measurement of P-selectin level in plasma (male=11, female=19). **g-h**. Flow cytometric analysis of multiple platelet-associated markers at the time points indicated. Total reactive oxygen species (ROS) levels in platelets (male=5, female=5); JC-1: ΔΨm depolarization of mitochondria (male=5, female=5); i. Flow cytometric analysis of apoptosis. Percentage of Annexin V positive platelets (male=5, female=5). j. ELISA measurement of thrombopoietin (TPO) level in plasma (male=11, female=20). **k**. Plasma proteomic analysis indicating a reduction in platelet count. Left panel, transition diagram of plasma proteomics. Shown in the diagram are the expression ratios for platelet formation-related proteins at the testing days compared with wFD1; middle panel, significant reduction in G6B and MYL9 shown by heatmap of G6B and MYL9. The color represents the level of protein expression at the indicated testing days; right panel, q-value for significant differences in protein accumulation. The X-axis represents a comparison between wFD7 and wFD1, and the Y-axis represents comparison between rFD7 and wFD1. Data are means ± SEM. \**P* < 0.05; \*\**P* < 0.01; \*\*\**P* < 0.001. *significant compared with wFD1, immediately before the start of fasting.

Platelet hematocrit (PCT) measures the volume occupied by platelets in the blood as a percentage. PCT was progressively reduced in fasting and refeeding (Figure 4b), results consistent with the observed reduction in platelet production. A mean platelet volume (MPV) test measures the average size of platelets. Beego did not lead to a significant change in MPV (Figure 4c). However, we did detect a progressive increase in platelet distribution width (PDW), indicating that beego increases the extent of platelet size variation (Figure 4d). A major function of platelets is their capacity for aggregation, which secures the ability for thrombus formation. We detected no beego-induced alteration of aggregation capacity, which was stable during fasting and refeeding (Figure 4e).

P-selectin is a cell adhesion protein that is mainly expressed on the membrane of activated vascular endothelial cells and activated thrombocytes, and the level of P-selectin on activated platelets reflects platelet activation. Our results show that platelet-localized P-selectin levels were significantly reduced during fasting and refeeding (Figure 4f). Beego with 14-day fasting and 7-day refeeding displayed similar patterns for the dynamic changes in the above platelet-related parameters (Supplementary Figure S4a-f).

To further examine if beego may affect platelet aging, we measured oxidative stress in platelets. Strikingly, reactive oxygen species (ROS) levels were significantly reduced during fasting, and this reduction was sustained after refeeding (Figure 4g), suggesting that beego confers a significant reduction in platelet oxidative stress. JC-1 assays measure Ψm depolarization of the mitochondria membrane, and such depolarization indicates active cell apoptosis and accelerated aging^59-61^. Beego significantly reduced the percentage of JC-1 green fluorescence (Figure 4h), suggesting an increase mitochondrial in mitochondrial membrane potential and thus a reduction in intrinsic pathway triggered activation of apoptosis. Consistently, flow cytometry analysis of Annexin V levels revealed a detectable beego-induced reduction in the extent of platelet apoptosis (Figure 4i). Thus, our data support the possibility that the observed reduction in platelet count in the normal platelet group may be caused by reduced overall production of platelets, rather than by increased apoptosis or aging. Beego with 14-day fasting and 7-day refeeding displayed a roughly similar pattern in the dynamic change of these parameters (Supplementary Figure S4g-i). These results suggest that in addition to limiting platelet generation (reducing thrombosis risk) without impairing hemostasis capacity, beego may also exert some effect in rejuvenating platelets.

To explore possible mechanisms by which beego reduces circulating platelet count (Figure 4a), we examined TPO level produced in the liver. Beego did not result in alteration of the TPO level (Figure 4j, Supplementary Figure S4j), suggesting that the observed reduction in platelet formation is not caused by regulatory molecules upstream of TPO expression. We therefore performed a proteomics analysis of platelets which revealed that, among the eight proteins essential for platelet formation, G6B and MYL9 were significantly reduced during fasting, and the levels of these two proteins remained low in refeeding (Figure 4k, left). These findings indicated that reduction of G6B and MYL9 may be responsible for the observed reduction of platelets, an idea supported further by our heatmap and Q-value analyses showing obvious and significant reductions in the G6B and MYL9 levels on days wFD7 and rFD7 as compared with wFD1 (Figure 4K, middle and right panels).

## Discussion

The present study is the first attempt describing the biological effects of beego, focusing on thrombosis and hemostasis. The results suggest that medically supervised beego for 7 or 14-day complete water-only fasting followed by 7-day refeeding, added with psychological intervention and breathing training as well as light excises, eventually reduce risks in thrombosis without compromising hemostasis.

A major feature of beego is its use of psychological induction or mind regulation to facilitate water-only fasting. Meditation is a critical traditional component in mind regulation during beego, and recent studies have confirmed positive effects of meditation on health^62-64^.

The glucose-to-ketone switch known to occur with some forms of incomplete fasting has been shown to lead to enhanced performance in cognition and mood, as well as motor and autonomic nervous system function^65,66^. Associated mechanisms for this switching in animal models involve the release of BDNF, a protein associated with neurogenesis and neuron protection^67-69^. However, there is to date no information about how BDNF or other biomolecules may contribute to health benefits obtained with human beego. Thus, it would be interesting to explore the connection between brain health and brain metabolic patterns during fasting^70^. In particular, we speculate it would be informative to determine if REST and FOXO1, two recently identified longevity-associated transcription factors^71^, may be involved in modulating neural excitation during meditation.

Deep breathing is also an important element in mind regulation during beego. It is traditionally believed that beego practitioners can obtain energy from nature to support their energy needs during fasting. This ideation may somehow spiritually support the practitioners, who had no scientific training or metabolic understanding to maintain their confidence during the fasting phase. Although such ideation appears to be scientifically baseless, deep breathing may help maintain sufficient body oxygen supply and body fluid pH value because accumulation of CO_2_ may reduce pH value in body fluid^1,2,72,73^. Notably, subjects were trained with two practices to use when they felt hunger: an air pharynx practice for “swallowing air” to imitate the action of food pharynx, as well as instructions for the beego subjects to repeatedly tell themselves “I have had a meal”. Although beego professionals in China believe the psychological induction and these practices are helpful for overcoming the feeling of possible hunger, in the future it will be imperative to explore the biological and psychological basis for mind regulation in beego.

While an overwhelmingly long list of studies have reported benefits and mechanisms for incomplete fasting that covering calorie restriction, intermittent fasting, fasting-mimicking diets, and periodic short-term water-only fasting with minimal calories or supplements^11^, studies of complete water-only fasting in human are much less numerous^3-10^. In our study, the changes in the lipid profile by beego are largely in concordance with the western styled fasting regimes^11,50,74,75^. Beego immediately results in significant triacylglycerol reduction selectively in triacylglycerol high subjects (Figure 1b), which is beneficial to reducing thrombosis risks.

It was notable that we found beego results in a transient increase in cholesterol levels during the fasting period of our beego program. An increase in the cholesterol level would typically be interpreted as a thrombosis risk. Beego with a fasting period of 7-14 days apparently causes opposing effects on metabolic dynamics between triacylglycerol and cholesterol. It was thus highly notable that in our study the increased cholesterol level (specifically LDL-C) in fasting was decreased in the refeeding period (Figure 1). The increase in blood cholesterol level may be caused in part by remobilization of cholesterol from adipose tissue during fasting^76,77^, or possibly by increased synthesis of cholesterol in liver to maintain the function of membrane system due to zero intake of dietary cholesterol during fasting^78,79^. An early report on complete starvation by six normal adults indicates a slight but non-significant change in cholesterol level^5^. Recent studies on cholesterol appear to have challenged the previous dogma that high level of this lipid molecule is a major risk factor for thrombosis. For example, one study indicates that high LDL-C is inversely associated with mortality in most people over 60 years, and that elderly people with high LDL-C live as long as or longer than those with low LDL-C^80^. Such accumulated findings have resulted in the modification of the US dietary guidelines in which no specific limit on daily dietary cholesterol is specified. Therefore, the transient LDL cholesterol increase in fasting may not necessarily be a significant risk factor. In fact, cholesterol is essential for the integrity of membrane functions^78,79^.

Abnormal activation of platelets is also a major cause for thrombosis^57,58^. Plasma proteomic analysis disclosed that beego attenuates platelet activation, aggregation and degranulation on fasting day 7 and refeeding day 7. The downregulation of the three aspects of platelet behaviors all contribute to the reduction of thrombosis risk. Similar to chronic 1-week subtotal fasting (<300 kcal/day as carbohydrates) that has mild anticoagulative, fibrinolytic and antiplatelet effect^75^ and to other chronic incomplete fasting regimes that decrease cardiovascular risk^81,82^, proteomic evidence in our study indicates that procoagulation and anticoagulation systems maintained unchanged by beego (Figure 2).

Intriguingly, circulating platelets appear to be rejuvenated after beego, as evidenced by beego-triggered reduction of aging markers in platelets (Figure 4g-i). For context, oxidative stress and mitochondrial membrane potential are both hallmarks of aging^30^, and loss of cells through apoptosis is also representative of aging; such cell loss can be regulated by caloric restriction^83^. The effect of the deceleration of platelet aging is consistent with better controlled activation of platelets with reduced P-selectin level (Figure 4f). Functional analysis of hearts and vessels also shows that some of the cardiovascular function were improved and rejuvenated. All of the benefits above are contributable to the reduction of cardiovascular risks. Importantly, these beneficial effects sustain when fasting is complete.

Surprisingly, our study showed that platelet count was reduced during beego (Figure 4a, left). Previous compete fasting studies did not measure platelets^3-10^, and incomplete fasting studies did not detect any changes in peripheral platelet numbers^11,50,74,75^. Given the facts that platelet count is decreased and hemostasis capacity remains unchanged, it suggests that platelet function is improved with beego. Since activation, apoptosis and aging of platelets were downregulated in beego (Figure 4f-i), reduction of platelet is not caused by shorter lifespan of the platelet, but possibly by an inhibition of platelet production in beego. Given the increase in blood cholesterol level, the body may react to reduce the production of platelet to limit thrombosis. Therefore, the biological significance for platelet reduction is presumably to reduce thrombosis risk. This contention is supported by the observation that beego did not reduce platelet counts in the platelet low subjects (Figure 4a, right). When platelet pool is insufficient to pose risk for thrombosis, beego will not reduce platelet formation. The contention that the reduction of palate counts is not caused by shorter lifespan is further supported by the observation that the reduction of platelet production is associated with significant reduction of MYL9 and G6B (Figure 4k). The MYL9 is the myosin light chain and implicated in megakaryopoiesis^84^, whereas G6B is a platelet receptor containing an immunoreceptor tyrosine-based inhibition motif, critically required in the formation of platelet^85,86^.

Apart from difficulty in validation of human beego effects with model animals due to the impossibility in psychological intervention with animals, a weakness of this study is that it is quite difficult to recruit a control group for comparison. The subjects voluntarily participated in the beego are either subhealthy individuals or individuals with metabolic syndrome. Some of them are on medication for controlling hypertension or diabetes before joining beego. The subjects who were on medication for hypertension or diabetes discontinued the medication in the entire course of the beego program. In our cohort, no life-threatening event or severe adverse effects occurred. Discontinuation of medication in beego is traditional and primarily empiric. While cautious evaluation is desired in the future, the safety of this practice is supported by complete fasting study led by the groups of Goldhamer and Compbell^7,8^. However, it may be risky for those individuals in medication to stop medication to serve as a control group for our study. It is also difficult to recruit sufficient healthy individuals to serve a control group to donate blood samples for multiple times for laboratory testing.

In conclusion, supervised beego eventually reduces thrombosis risk without compromising hemostasis capacity. The risk in beego is mild and reversible on the completion of this practice, and the benefits to cardiovascular health are significant, efficient and sustainable (Figure 5). Therefore, beego can serve as an alternative non-invasive intervention on those with metabolic syndrome or in overnutritional conditions who meet the inclusion criteria detailed in the methods. At minimum, our work supports that future studies are warranted to determine suitable frequencies for beego practice for individuals with specific health needs, to determine if use of cholesterol-lowering drugs is necessary in the fasting period of the beego program, and to further explore mechanistic basis of the multiple physiological changes we observed during fasting and refeeding. Hopefully, this study will motivate more researchers to join us in the pursuit and improvement of the health efficacy of this traditional Chinese practice in a modern context.

**Figure 5.**
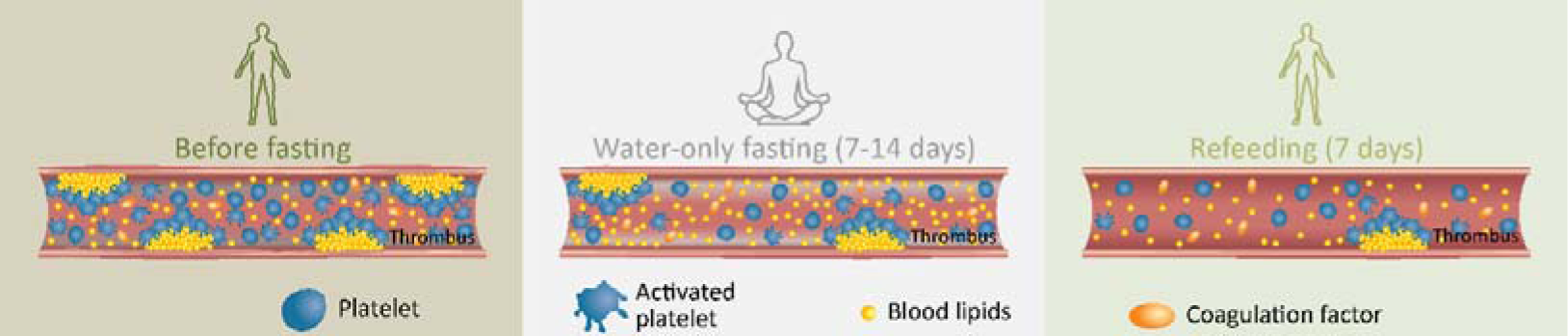
Graphical summary on the effects of beego on thrombosis and hemostasis. In a beego program, the potentially transit mild thrombosis risk in fasting period is reversible by refeeding; Beego maintains hemostasis capacity and limits platelet generation and activation to reduce thrombosis risk.

## Methods

### Ethics

The study was conducted with approval from Institutional Ethics Review Board at Soochow University (Approval No. ECSU-2019000153) and the Chinese Clinical Trial Register, an official review board for clinical trial (Registration No. ChiCTR1900027451), affiliated to The Ministry of Public Health of China (http://www.chictr.org.cn/index.aspx). All participants were recruited by the Soyo Center of Soochow University, an institutional platform for studying non-medical intervention for fitness. Participants were enrolled after giving their written informed consent. Thirty-eight participants aged from 27 to 67 years were enrolled.

### Exclusion criteria

Participants met eligibility criteria and had no predefined contraindication to beego practice. The exclusion criteria included, but were not limited to the following:

1) Acute diseases, active tuberculosis, gastrointestinal ulcer with gastrointestinal bleeding; 2) significant lesions in the organs, including advanced kidney, liver, or cerebrovascular insufficiency; 3) thin and weak patients with chronic hepatitis, thin and weak patients with diabetes, diabetes patients with injection of insulin for more than five successive years, nephropathy complicated with hypertension for more than ten years, intense valvular heart disease or severe cardiac arrhythmias, second or third stage lung gangrene patients; 4) advanced cancer patients, or any stage of pancreatic cancer, or thin and weak cancer patients; 5) individuals with a history of organ transplantation, or frequent organ bleeding; 6) thyropathy in long-term medication; 7) individuals with severe active allergic symptoms except pollen allergy; 8) cachexia individuals with body weight below 40 kg, or loss of hearing and loss of vision; 9) infectious diseases, excluding cold; 10) certain inherited disorders such as medium-chain acyl-CoA dehydrogenase deficiency and porphyria; 11) dementia or other severely debilitating cognitive disease, severe depression, hysteria, epilepsy, other in critical care patients; 12) existing pregnancy, lactation period, or age above 70 and below 20.

### Beego program

The beego program was supervised by a combined group of certified professionals in medicine, life sciences, health management, and nursing for physical examination. Emergency rescue kits were provided during the critical period of first 4-day fasting. Beego in this study comprised a period of water-only fasting for 7 or 14 days, followed by a refeeding period for another 7 days with smooth and gradual incensement of nutrients. The subjects with subhealthy conditions (e.g., overweight, metabolic syndrome) participated voluntarily in the beego program for preventive or therapeutic reasons. Medication (for hypertension or diabetes) was not taken during the entire period of the beego, largely based on previous studies^7,8,87^ as well as our previous practice. 38 began the beego intervention, among which 31 subjects (11 male and 20 female) chose 7-day fasting and 7-day refeeding program, while 7 subjects (3 male, 4 female) chose 14-day fasting and 7-day refeeding program. After beginning of the beego, the participants were given water but no food; participates could drink water *ad libitum*.

To facilitate the complete water-only fasting, psychological induction (i.e., a mind regulation program), including but not limited to meditation, was conducted with the subjects over the first four days of the fasting period. Specifically, at the beginning of the fasting period, the subjects, in a sitting meditation position, participated in a deep breathing program (abdominal breath) to ensure that they inhaled adequate oxygen and exhaled carbon dioxide^1,2^ as follows: abdominal breathing moves the diaphragm up and down. The diaphragm drops upon inhalation, squeezing the organs below, so the stomach expands instead of the chest. Upon exhalation, the diaphragm rises higher than usual, enabling deep breathing which promotes exhalation of the carbon dioxide that tends to stagnate at the bottom of the lungs^72,73^. Subjects assume a supine or comfortable sitting position, put one hand on the belly button of the abdomen to relax, and breathe naturally, while seeking to inhale to maximize the expansion of the abdomen to make the abdomen bulge and keep the chest motionless. When exhaling, the abdomen naturally recesses and draws inward toward the spine, while the chest remains motionless, maximizing the inward contraction of the abdomen and exhaling all of the exhaust gas from the lungs. Each round of meditation along with abdominal breath lasted about 45 min, followed by light physical exercise for 3-5 min (including for example self-tapping on their bodies and stretching arms and legs to make the body feel comfortable). During the fasting program, the beego subjects were instructed to conduct self-talk by saying “I have had a meal” and were trained in a “swallowing practice” for air pharynx when they felt hungry.

At the designated location, subjects fasted together for the first four days; subsequently, they fasted on their own at their local home or workplace. Beginning on fasting day 5, participates were able to resume office work or other easy work tasks (excluding tough physical endeavor). During the fasting period, the subjects were counseled to arrange their work schedules to be staggered with regular mealtimes (to reduce the acquired reflex for meal). Manual labor or vigorous exercise was avoided throughout the beego program. Water-only fasting was discontinued when any participant decided to give up the fasting or when the licensed professionals in health management deemed it necessary for medical reasons.

### Refeeding Protocol

Following the water-only fasting, subjects refed for a period of time lasting no less of half of the fasting length. We specified a particular diet and eating protocol for each of 7 refeeding days. An example is given below for day 1, 2 and for day 5, with the remaining details available in the supplementary information. Note that ingredient availability and typical kitchen implements are quite homogenous for most Chinese households, and counselors worked with the participants to customize serving sizes.

### --Refeeding day 1 and 2

Morning, midday, and evening: prepare a rice soup cooked with a small amount of millet by boiling the rice thoroughly with water. Only the liquid soup should be consumed (i.e., do not eat rice grains, consume no more than half a bowl of this soup at a given feeding session, but feel free to consume this soup as many as 6 or 7 times on day 1 and day 2.

### --Refeeding day 5

For breakfast, you may have gruel or boiled noodles, with small amount of oil and salt; you may eat boiled eggs, but only eat egg yolk. For noon meal, you may eat a small amount of rice, and rice should be soft, and eat fried green vegetables with less oil and salt. it is best to drink gruel in the evening. On the fifth day of reeating, you may add an appropriate amount of fruits, such as pears, watermelons and other fruits with more water. Don’t eat too much.

### Sample collection

To document the effect of beego on health, we performed the following predefined measurements: Blood samples were collected at 7:00 to 9:00 am of the testing days, with group one on fasting day 1, 4, 7 and refeeding day 7, and another group on fasting day 1,4,7,14 and refeeding day 7. The samples were collected by venipuncture through a safety needle into an blood collection tube containing the anticoagulation sodium citrate (final concentration of 0.32% by weight, 1:9 ratio of citrate to blood), previously established to minimize preanalytical variables in platelet function testing. Note that we also collected data at baseline (just before fasting on wFD1). The information of reagents used in lab tests is given in Supplementary Table 1.

### Blood pressure and pulse wave

Blood pressure was measured after a pause, once at the nondominant arm in a sitting position (upper arm blood pressure monitor, OMRON). Pulse wave was monitored by Mobil-O-Graph PWA (Germany) according to the manufacturer’s protocols (AL Rowais Medical Equipment).

### Blood lipid tests

The tests for blood lipids were measured just before fasting commencement, during fasting, and refeeding in the morning between 7:00–9:00 am. Whole blood without anticoagulant was incubated at 4°C overnight, and then centrifuged at 500 g for 20 min at room temperature. All serum samples were separated and frozen at −80°C until laboratory testing. Serum was analyzed for blood lipids with the Chemistry Analyzer (Roche, cobas c702) by Dian Diagnostics, China.

### Coagulation tests

A medical professional took a blood sample and send it to our hospital for testing and analysis. All assays were performed on an automated coagulation analyzer (Sysmex, cs-2000i) with corresponding reagents (Siemens). The tests included prothrombin times (PTs), internal normalized ratio (INR), activated partial thromboplastin times (APTTs), fibrinogen (FIB), antithrombin III (AT III), and thrombin time (TT). These tests were conducted according to the standard operating procedures (SOP) of the instrument.

### Complete blood count

The samples were analyzed on the Automatic Five Categories Hematology Analyzer (Siemens) on which were observed platelet indices including platelet (PLT) count, mean platelet volume (MPV), platelet distribution width (PDW), and plateletcrit (PCT).

### Platelet preparation

The collected whole blood with the anticoagulant was centrifuged at 200 x g for 20 min at room temperature (RT) with no brake applied to prevent any platelet activation. The supernatant platelet-rich plasma (PRP) must then gently transferred to a fresh tube using a polypropylene Pasteur pipette and the PRP is stored at RT. The prepared PRP was mixed with 1 μM PGE-1(ENZO). Washed platelets was collected from PRP by 500 x g centrifugation for 20 minutes at RT and resuspended in modified Tyrode’s Buffer containing 137 mM NaCl, 20 mM HEPES, 1 mM MgCl_2_·6H_2_O, 2.7 mM KCl, 3.3 mM Na_2_HPO_4_·2H_2_O, 5.6 mM D-glucose and 1g/L BSA (pH 7.4).The supernatant plasma was packed, and then stored at −80°C.

### Platelet aggregation

Washed platelets were normalized to a final concentration of 2-4 × 10^8^ /mL with Tyrode’s Buffer. Diluted washed platelet (250 μL) was placed in an aggregometer cuvette, warmed to 37°C and measured in a Dual-channel platelet aggregation apparatus (Chrono-log). After 1 mM CaCl_2_ was added, baseline measurements were obtained, 0.012 U/mL thrombin (Chrono-log) was added to induce aggregation. The percent aggregation was calculated from the amplitude of the tracings at 5 minutes and normalized to the response of the untreated control within an individual experiment.

### Soluble P-selectin in plasma

Plasma soluble P-selectin concentrations were assayed using a Human P-Selectin ELISA Kit (Affandi) on Labsystem Multiskan MS (Thermo) according to the manufacturer’s instructions.

### Thrombopoietin

Thrombopoietin levels in plasma were assayed by enzyme-linked immunosorbent assay (ELISA) using a Human Thrombopoietin, TPO ELISA Kit (Affandi) according to the manufacturers’ instructions (eBioscience).

### Phosphatidylserine externalization assay

The prepared PRP (1-3×10^6^ cells) was labeled with 5 μL Mouse Anti-Human CD41a (BD Biosciences) at 37°C for 30 minutes in the dark, then incubated with CaCl_2_ (1 mM) and thrombin (0.2 U/mL) at RT for 15 minutes. The pretreated PRP was mixed with 5 μL diluted Annexin V–FITC (BD Biosciences). Samples were gently mixed and incubated at RT in the dark for 20 minutes, diluted with 150 μL Annexin V–binding buffer, and analyzed using a flow cytometer (Backman Gallious).

### Mitochondrial ΔΨ m depolarization assay

Washed platelet (1-3×10^6^ cells) was incubated with CaCl_2_ (1 mM) and thrombin (0.02 U/mL) at RT for 5 minutes. The pretreated platelets were incubated with 2 μg/mL JC-1 antibody (Beyotime) for 30 minutes at 37°C, and then diluted with 150 μL Tyrode’s buffer. The treated samples were detected by flow cytometry.

### Oxidative stress level detection

Washed platelet (1-3×10^6^ cells) was labeled with 5 μL Mouse Anti-Human CD41a (BD Biosciences) at 37°C for 30 minutes then incubated with CaCl_2_ (1 mM) and thrombin (0.02 U/mL) at RT for 5 minutes. Treated platelets were incubated with 10 μM CM-H2DCFDA (Thermo Fisher Scientific) for 30 minutes at 37°C, and then diluted with 150 μL Tyrode’s buffer. The treated samples were detected by flow cytometry.

### Flow cytometry

Flow cytometric analysis of platelets was conducted with Beckman Coulter (Gallios) as described previously^59,88^.

### Plasma proteomics

#### Sample preparation

SDS free lysate was added to 100 μL serum/plasma sample to make up a total volume of 1 mL; DTT was added to sample with a final concentration of 10 mM; samples were incubated at 37°C for 30 min; Iodoacetamide was added to sample with a final concentration of 55 mM, and incubated in the dark at room temperature for 30 min. The mixture of proteins was passed through a solid phase extraction (SPE) C18 column for protein enrichment. The sample protein concentration was calculated based on the standard curve and sample OD595, then performed SDS-PAGE. Proteins were hydrolyzed by trypsin enzyme and the enzymatic peptides were desalted using a Strata X column and vacuumed to dryness. **DIA analysis by nano-LC-MS/MS**. The peptides separated by liquid phase chromatography (LC-MS/MS) were ionized by a nanoESI source and then passed to a tandem mass spectrometer Q-Exactive HF X (Thermo Fisher Scientific, San Jose, CA) for DDA (data-dependent acquisition) mode detection. The peptides separated by liquid phase chromatography were ionized by a nanoESI source and passed to a tandem mass spectrometer Q-Exactive HF X (Thermo Fisher Scientific, San Jose, CA) for DIA (data-independent acquisition) mode detection.

### Bioinformatic Analysis Pipeline

This process is based on the sample data generated from a high-resolution mass spectrometer. DDA data was identified by Andromeda search engine within MaxQuant, and Spectronaut™ used identification results for spectral library construction. For large-scale DIA data, Spectronaut™ uses the constructed spectral library information to complete deconvolution and extraction, and uses the mProphet algorithm to complete analytical quality control, thus obtaining a large number of reliable quantitative results. This pipeline also performed GO, COGs, Pathway functional annotation analysis and time series analysis. Based on the quantitative results, the differential proteins between comparison groups were found, and finally function enrichment analysis, protein-protein interaction (PPI) and subcellular localization analysis of the differential proteins were performed.

### Statistical analyses

Laboratory technicians who performed the measurements and data statistics were blind to the information of the samples collected before, during, and after the intervention. Statistical analyses were performed using SPSS version 22.0. Differences between groups were analyzed using univariate repeated-measures analysis of variance (RM ANOVA). To assess for changes during fasting, we performed each fasting time point to baseline (wFD1) using One-way analysis of variance ^18^ with multiple comparisons. Multiple comparisons were performed using Dunnett’s test and then using the Benjamini & Hochberg method to control the false discovery rate. Data is expressed as mean ± standard error of the mean (SEM), which is provided in the Supplementary Information (Supplementary Table S1, S2). *P*<0.05 was considered to indicate a statistically significant difference.

## Data Availability

The data that support the findings of this study are available from the corresponding authors upon request.

## Abbreviations

AIx: augmentation index
APTT: activated partial thromboplastin time
AT III: antithrombin III testing
BDNF: brain-derived neurotrophic factor
CNS: central nervous system
COGs: cluster of orthologous groups
DBP: diastolic blood pressure
DDA: data-dependent acquisition
DIA: data-independent acquisition
DTT: dithiothreitol
ELISA: Enzyme-linked immunosorbent assay
FIB: Fibrinogen
FITC: fluoresceine isothiocyanate
GO: Gene ontology
HDL-C: high-density lipoprotein cholesterol
INR: international normalized ratio
LC-MS/MS: liquid phase chromatography
LDL-C: low-density lipoprotein cholesterol
MPV: Mean platelet volume
PCT: platelet hematocrit
PDW: platelet distribution width
PE: phycoerythrin
PLT: platelet
PPI: protein-protein interaction
PRP: platelet-rich plasma
PT: prothrombin time
PWA: pulse wave
PWV: pulse wave velocity
rFD: refeeding days
RM: repeated measures
ROS: reactive oxygen species
RT: room temperature
SBP: systolic blood pressure
SEM: standard error of the mean
SOP: standard operating procedures
SPE: solid phase extraction
TC: total cholesterol
TG: triacylglycerol
TPO: thrombopoietin
TT: thrombin time
wFD: water-only fasting day.

## Acknowledgments

We thank all the volunteers participating in this beego study and the campus Hospital of Soochow University for providing the facilities in the collection of blood samples and taking physical measurement. We further thank Mr. Yaozhong Wu and his team for their advice on the beego protocol and Dr. Quansheng Zhou and Dr. Yun Zhao for their critical reading of the manuscript. This study was supported in part by the National Natural Science Foundation of China by grants 91649113 and 31771640 (to JW), 82000117 (to YF), and by Soyo Center of Soochow University (to JW) by grant H190443, and by the Natural Science Foundation of Jiangsu Province, China (to YF) by grant BK20200191, and by The Postdoctor Science Foundation of Jiangsu Province (to YF) by grant 2020Z064.

## Author contributions

Y.F. and J.W. conceived and planned the study. J.W., Y.F., N.Y. and S.Z. recruited the participate cohort. Y.G., L.Z., C.Z. and Y.L collected tissue biopsies. Y.F. and J.Q. conducted proteomic analysis of the plasma. Y.F., Y.G. and C.Z. generated the figures. Y.F and J.W. analyzed the data and wrote the manuscript. N.Y., S.Z., L.W., M.L., L.X., W.W., L.L., L.J., X.G., J.Z., Z.C., Y.S., G.W. and K.D. contributed to data analysis or discussion. All authors read and agreed upon the submitted manuscript.

## Competing interests

The authors declare no competing interests.

## Additional information

Extended data is included in the Supplementary Information file.

## Notes

### Competing Interest Statement

The authors have declared no competing interest.

### Clinical Trial

http://www.chictr.org.cn/index.aspx, number, ChiCTR1900027451

### Funding Statement

This study was supported by the National Natural Science Foundation of China (to JW) by grants 91649113 and 31771640, and by Soochow University (to JW) by grant H190443, and by the Natural Science Foundation of Jiangsu Province, China (to YF) by grant SBK2020043689, and by The Postdoctor Science Foundation of Jiangsu Province (to YF) by grant 2020Z064, and by the Priority Academic Program Development of Jiangsu Higher Education Institutions, Jiangsu Province, China.

### Author Declarations

Approval No. ECSU-2019000153 by Ethics Review Board at Soochow University, China

### Summary of Updates

Beego is a traditional Chinese complete water-only fasting practice initially developed for spiritual purposes, later extending to physical fitness purposes. Beego notably includes a psychological induction component that includes meditation and abdominal breathing, light body exercise, and ends with a specific gradual refeeding program before returning to a normal diet. Beego has regained its popularity in recent decades in China as a strategy for helping people in subhealthy conditions or with metabolic syndrome, but we are unaware of any studies examining the biological effects of this practice. To address this, we here performed a longitudinal study of beego comprising fasting (7 and 14 day cohorts) and a 7-day programmed refeeding phase. In addition to detecting improvements in cardiovascular physiology and selective reduction of blood pressure in hypertensive subjects, we observed that beego decreased blood triacylglycerol (TG) selectively in TG-high subjects and increased cholesterol in all subjects during fasting; however, the cholesterol levels were normalized after completion of the refeeding program. Strikingly, beego reduced platelet formation, activation, aggregation, and degranulation, resulting in an alleviated thrombosis risk, yet maintained hemostasis by sustaining levels of coagulation factors and other hemostatic proteins. Mechanistically, we speculate that downregulation of G6B and MYL9 may influence the observed beego-mediated reduction in platelets. Fundamentally, our study supports that supervised beego reduces thrombosis risk without compromising hemostasis capacity. Moreover, our results support that beego under medical supervision can be implemented as noninvasive intervention for reducing thrombosis risk, and suggest several lines of intriguing inquiry for future studies about this fasting practice (http://www.chictr.org.cn/index.aspx, number, ChiCTR1900027451).

## References

1. Wei, G.X., et al. Tai Chi Chuan modulates heart rate variability during abdominal breathing in elderly adults. Psych Jn 5, 69–77 (2016).

2. Byeon, K., et al. The response of the vena cava to abdominal breathing. J Altern Complement Med 18, 153–157 (2012).

3. Bloom, W.L. Fasting as an introduction to the treatment of obesity. Metabolism 8, 214–220 (1959).

4. Drenick, E.J., Swendseid, M.E., Blahd, W.H. & Tuttle, S.G. Prolonged Starvation as Treatment for Severe Obesity. JAMA 187, 100–105 (1964).

5. Consolazio, C.F., et al. Metabolic aspects of acute starvation in normal humans: performance and cardiovascular evaluation. Am J Clin Nutr 20, 684–693 (1967).

6. Benedict, F.G. A study of prolonged fasting / by Francis Gano Benedict, (Carnegie Institution of Washington, Washington, 1915).

7. Goldhamer, A., Lisle, D., Parpia, B., Anderson, S.V. & Campbell, T.C. Medically supervised water-only fasting in the treatment of hypertension. J Manipulative Physiol Ther 24, 335–339 (2001).

8. Ciurleo, A. & Marchese, M. Medically supervised water-only fasting in the treatment of hypertension. J Manipulative Physiol Ther 25, 138-139; author reply 139 (2002).

9. Finnell, J.S., Saul, B.C., Goldhamer, A.C. & Myers, T.R. Is fasting safe? A chart review of adverse events during medically supervised, water-only fasting. BMC Complement Altern Med 18, 67 (2018).

10. Lategola, M. EFFECT OF 5-DAY COMPLETE STARVATION ON CARDIOPULMONARY FUNCTIONS OF AEROBIC WORK CAPACITY AND ORTHOSTATIC TOLERANCE. in Federation Proceedings, Vol. 24 590-& (FEDERATION AMER SOC EXP BIOL 9650 ROCKVILLE PIKE, BETHESDA, MD 20814–3998, 1965).

11. de Cabo, R. & Mattson, M.P. Effects of Intermittent Fasting on Health, Aging, and Disease. N Engl J Med 381, 2541–2551 (2019).

12. Steinhauser, M.L., et al. The circulating metabolome of human starvation. JCI Insight 3(2018). 13.

13. Capozzi, M.E., et al. The Limited Role of Glucagon for Ketogenesis During Fasting or in Response to SGLT2 Inhibition. Diabetes 69, 882-892 (2020).

14. Lopez-Soldado, I., Bertini, A., Adrover, A., Duran, J. & Guinovart, J.J. Maintenance of liver glycogen during long-term fasting preserves energy state in mice. FEBS Lett 594, 1698–1710 (2020).

15. Volek, J.S., et al. Body composition and hormonal responses to a carbohydrate-restricted diet. Metabolism 51, 864–870 (2002).

16. Fontana, L., et al. Effects of 2-year calorie restriction on circulating levels of IGF-1, IGF-binding proteins and cortisol in nonobese men and women: a randomized clinical trial. Aging Cell 15, 22–27 (2016).

17. Sutton, E.F., et al. Early Time-Restricted Feeding Improves Insulin Sensitivity, Blood Pressure, and Oxidative Stress Even without Weight Loss in Men with Prediabetes. Cell Metab 27, 1212–1221 e1213 (2018).

18. Jordan, S., et al. Dietary Intake Regulates the Circulating Inflammatory Monocyte Pool. Cell 178, 1102–1114 e1117 (2019).

19. Collins, N., et al. The Bone Marrow Protects and Optimizes Immunological Memory during Dietary Restriction. Cell 178, 1088–1101 e1015 (2019).

20. Choi, I.Y., et al. A Diet Mimicking Fasting Promotes Regeneration and Reduces Autoimmunity and Multiple Sclerosis Symptoms. Cell Rep 15, 2136–2146 (2016).

21. Cignarella, F., et al. Intermittent Fasting Confers Protection in CNS Autoimmunity by Altering the Gut Microbiota. Cell Metab 27, 1222–1235 e1226 (2018).

22. Nagai, M., et al. Fasting-Refeeding Impacts Immune Cell Dynamics and Mucosal Immune Responses. Cell 178, 1072–1087 e1014 (2019).

23. Mladenovic Djordjevic, A., Loncarevic-Vasiljkovic, N. & Gonos, E.S. Dietary restriction and oxidative stress: friends or enemies? Antioxid Redox Signal (2020).

24. Kong, J., et al. Spatiotemporal contact between peroxisomes and lipid droplets regulates fasting-induced lipolysis via PEX5. Nat Commun 11, 578 (2020).

25. Longo, V.D. & Mattson, M.P. Fasting: molecular mechanisms and clinical applications. Cell Metab 19, 181–192 (2014).

26. Liu, Y., et al. Short-term caloric restriction exerts neuroprotective effects following mild traumatic brain injury by promoting autophagy and inhibiting astrocyte activation. Behav Brain Res 331, 135–142 (2017).

27. Colman, R.J., et al. Caloric restriction delays disease onset and mortality in rhesus monkeys. Science 325, 201–204 (2009).

28. Mattison, J.A., et al. Impact of caloric restriction on health and survival in rhesus monkeys from the NIA study. Nature 489, 318–321 (2012).

29. Johnson, J.B., et al. Alternate day calorie restriction improves clinical findings and reduces markers of oxidative stress and inflammation in overweight adults with moderate asthma. Free Radic Biol Med 42, 665–674 (2007).

30. Redman, L.M., et al. Metabolic Slowing and Reduced Oxidative Damage with Sustained Caloric Restriction Support the Rate of Living and Oxidative Damage Theories of Aging. Cell Metab 27, 805–815 e804 (2018).

31. Horne, B.D., et al. Usefulness of routine periodic fasting to lower risk of coronary artery disease in patients undergoing coronary angiography. Am J Cardiol 102, 814–819 (2008).

32. Horne, B.D., et al. Randomized cross-over trial of short-term water-only fasting: metabolic and cardiovascular consequences. Nutr Metab Cardiovasc Dis 23, 1050–1057 (2013).

33. Most, J., et al. Significant improvement in cardiometabolic health in healthy nonobese individuals during caloric restriction-induced weight loss and weight loss maintenance. Am J Physiol Endocrinol Metab 314, E396–E405 (2018).

34. Ajona, D., et al. Short-term starvation reduces IGF-1 levels to sensitize lung tumors to PD-1 immune checkpoint blockade. 1, 75–85 (2020).

35. Yamaza, H., et al. FoxO1 is involved in the antineoplastic effect of calorie restriction. Aging Cell 9, 372–382 (2010).

36. Di Biase, S., et al. Fasting-Mimicking Diet Reduces HO-1 to Promote T Cell-Mediated Tumor Cytotoxicity. Cancer Cell 30, 136–146 (2016).

37. Pietrocola, F., et al. Caloric Restriction Mimetics Enhance Anticancer Immunosurveillance. Cancer Cell 30, 147–160 (2016).

38. Nencioni, A., Caffa, I., Cortellino, S. & Longo, V.D. Fasting and cancer: molecular mechanisms and clinical application. Nat Rev Cancer 18, 707–719 (2018).

39. Kjeldsen-Kragh, J., et al. Controlled trial of fasting and one-year vegetarian diet in rheumatoid arthritis. Lancet 338, 899–902 (1991).

40. Li, C., et al. Metabolic and psychological response to 7-day fasting in obese patients with and without metabolic syndrome. Forsch Komplementmed 20, 413–420 (2013).

41. Steiniger, J., Schneider, A., Bergmann, S., Boschmann, M. & Janietz, K. [Effects of fasting and endurance training on energy metabolism and physical fitness in obese patients]. Forsch Komplementmed 16, 383–390 (2009).

42. Schmidt, S., et al. [Uncontrolled clinical study of the efficacy of ambulant fasting in patients with osteoarthritis]. Forsch Komplementmed 17, 87–94 (2010).

43. Michalsen, A., et al. In-Patient Treatment of Fibromyalgia: A Controlled Nonrandomized Comparison of Conventional Medicine versus Integrative Medicine including Fasting Therapy. Evid Based Complement Alternat Med 2013, 908610 (2013).

44. Michalsen, A., et al. Prolonged fasting in patients with chronic pain syndromes leads to late mood-enhancement not related to weight loss and fasting-induced leptin depletion. Nutr Neurosci 9, 195–200 (2006).

45. Witte, A.V., Fobker, M., Gellner, R., Knecht, S. & Floel, A. Caloric restriction improves memory in elderly humans. Proc Natl Acad Sci U S A 106, 1255–1260 (2009).

46. Gill, S., Le, H.D., Melkani, G.C. & Panda, S. Time-restricted feeding attenuates age-related cardiac decline in Drosophila. Science 347, 1265–1269 (2015).

47. Michalsen, A., et al. Incorporation of fasting therapy in an integrative medicine ward: evaluation of outcome, safety, and effects on lifestyle adherence in a large prospective cohort study. J Altern Complement Med 11, 601–607 (2005).

48. Martin, C.K., et al. Effect of Calorie Restriction on Mood, Quality of Life, Sleep, and Sexual Function in Healthy Nonobese Adults: The CALERIE 2 Randomized Clinical Trial. JAMA Intern Med 176, 743–752 (2016).

49. Trepanowski, J.F., et al. Effect of Alternate-Day Fasting on Weight Loss, Weight Maintenance, and Cardioprotection Among Metabolically Healthy Obese Adults: A Randomized Clinical Trial. JAMA Intern Med 177, 930–938 (2017).

50. Wilhelmi de Toledo, F., Grundler, F., Bergouignan, A., Drinda, S. & Michalsen, A. Safety, health improvement and well-being during a 4 to 21-day fasting period in an observational study including 1422 subjects. PLoS One 14, e0209353 (2019).

51. Abdel-Latif, A. & Smyth, S.S. Preventing platelet thrombosis with a PAR1 pepducin. Circulation 126, 13–15 (2012).

52. Langer, H.F., Weber, C. & Gawaz, M. The platelet--thrombosis and beyond. Thromb Haemost 110, 857–858 (2013).

53. Baron, J.A., et al. Gastrointestinal adverse effects of short-term aspirin use: a meta-analysis of published randomized controlled trials. Drugs R D 13, 9–16 (2013).

54. Born, G.V. Platelets and blood vessels. J Cardiovasc Pharmacol 6 Suppl 4, S706–713 (1984).

55. Gosseye, S., et al. Platelet aggregates in small lung vessels and death during liver transplantation. Lancet 338, 532–534 (1991).

56. Yougbare, I., et al. Maternal anti-platelet beta3 integrins impair angiogenesis and cause intracranial hemorrhage. J Clin Invest 125, 1545–1556 (2015).

57. Du, X. & Ginsberg, M.H. Signaling and platelet adhesion. in Advances in Molecular and Cell Biology, Vol. 28 269–301 (Elsevier,1999).

58. Li, Z., et al. A stimulatory role for cGMP-dependent protein kinase in platelet activation. Cell 112, 77–86 (2003).

59. Zhao, L., et al. Protein kinase A determines platelet life span and survival by regulating apoptosis. J Clin Invest 127, 4338–4351 (2017).

60. Chen, M., et al. Akt-mediated platelet apoptosis and its therapeutic implications in immune thrombocytopenia. Proc Natl Acad Sci U S A 115, E10682–E10691 (2018).

61. Vyssokikh, M.Y., et al. Mild depolarization of the inner mitochondrial membrane is a crucial component of an anti-aging program. Proc Natl Acad Sci U S A 117, 6491–6501 (2020).

62. Edwards, M.K. & Loprinzi, P.D. Comparative effects of meditation and exercise on physical and psychosocial health outcomes: a review of randomized controlled trials. Postgrad Med 130, 222–228 (2018).

63. Sinha, S.S., Jain, A.K., Tyagi, S., Gupta, S.K. & Mahajan, A.S. Effect of 6 Months of Meditation on Blood Sugar, Glycosylated Hemoglobin, and Insulin Levels in Patients of Coronary Artery Disease. Int J Yoga 11, 122–128 (2018).

64. Murphy, J.A., et al. Pilot-Testing of “Healthy Body Healthy Mind”: An Integrative Lifestyle Program for Patients With a Mental Illness and Co-morbid Metabolic Syndrome. Front Psychiatry 10, 91 (2019).

65. Cahill, G.F., Jr. & Veech, R.L. Ketoacids? Good medicine? Trans Am Clin Climatol Assoc 114, 149-161; discussion 162-143 (2003).

66. Mattson, M.P., Moehl, K., Ghena, N., Schmaedick, M. & Cheng, A. Intermittent metabolic switching, neuroplasticity and brain health. Nat Rev Neurosci 19, 63–80 (2018).

67. Mattson, M.P., Maudsley, S. & Martin, B. BDNF and 5-HT: a dynamic duo in age-related neuronal plasticity and neurodegenerative disorders. Trends Neurosci 27, 589–594 (2004).

68. Mattson, M.P. & Wan, R. Beneficial effects of intermittent fasting and caloric restriction on the cardiovascular and cerebrovascular systems. J Nutr Biochem 16, 129–137 (2005).

69. Marosi, K., et al. 3-Hydroxybutyrate regulates energy metabolism and induces BDNF expression in cerebral cortical neurons. J Neurochem 139, 769–781 (2016).

70. Owen, O.E., et al. Brain metabolism during fasting. J Clin Invest 46, 1589–1595 (1967).

71. Zullo, J.M., et al. Regulation of lifespan by neural excitation and REST. Nature 574, 359–364 (2019).

72. Kintner, E.P. Acid base, blood gas, and electrolyte balances. Prog Clin Pathol 4, 143–180 (1972).

73. Falke, K.J., et al. Breathing pattern, CO2 elimination and the absence of exhaled NO in freely diving Weddell seals. Respir Physiol Neurobiol 162, 85–92 (2008).

74. Vaisman, N., Sklan, D. & Dayan, Y. Effect of moderate semi-starvation on plasma lipids. Int J Obes 14, 989–996 (1990).

75. Huber, R., et al. Effects of subtotal fasting on plasmatic coagulation, fibrinolytic status and platelet activation: a controlled pilot study in healthy subjects. Nutr Metab Cardiovasc Dis 15, 212–218 (2005).

76. Tobey, J.A. The biology of human starvation. (American Public Health Association, 1951).

77. ENDE, N.J.T.A.J.o.C.N. Starvation Studies: With Special Reference to Cholesterol. 11, 270–280 (1962).

78. Chang, T.Y., Chang, C.C., Ohgami, N. & Yamauchi, Y. Cholesterol sensing, trafficking, and esterification. Annu Rev Cell Dev Biol 22, 129–157 (2006).

79. Chu, B.B., et al. Cholesterol transport through lysosome-peroxisome membrane contacts. Cell 161, 291–306 (2015).

80. Ravnskov, U., et al. Lack of an association or an inverse association between low-density-lipoprotein cholesterol and mortality in the elderly: a systematic review. BMJ Open 6, e010401 (2016).

81. Ridker, P.M., Buring, J.E. & Rifai, N. Soluble P-selectin and the risk of future cardiovascular events. Circulation 103, 491–495 (2001).

82. Hackam, D.G. & Anand, S.S. Emerging risk factors for atherosclerotic vascular disease: a critical review of the evidence. JAMA 290, 932–940 (2003).

83. Trembinski, D.J., et al. Aging-regulated anti-apoptotic long non-coding RNA Sarrah augments recovery from acute myocardial infarction. Nat Commun 11, 2039 (2020).

84. Gilles, L., et al. MAL/SRF complex is involved in platelet formation and megakaryocyte migration by regulating MYL9 (MLC2) and MMP9. Blood 114, 4221–4232 (2009).

85. Coxon, C.H., Geer, M.J. & Senis, Y.A. ITIM receptors: more than just inhibitors of platelet activation. Blood 129, 3407–3418 (2017).

86. Geer, M.J., et al. Uncoupling ITIM receptor G6b-B from tyrosine phosphatases Shp1 and Shp2 disrupts murine platelet homeostasis. Blood 132, 1413–1425 (2018).

87. Binsalih, S., Al Sayyari, R.A., Sheikho, M., Hejaili, F.F. & Al Sayyari, A.A. Effect of Fasting the Whole Month of Ramadan on Renal Function Among Muslim Patients With Kidney Transplant: A Meta-Analysis. Exp Clin Transplant 17, 588–593 (2019).

88. Mason, K.D., et al. Programmed anuclear cell death delimits platelet life span. Cell 128, 1173–1186 (2007).

