## Supplementary figures and tables for "The impact of supervised beego, a traditional Chinese water-only fasting, on thrombosis and hemostasis"

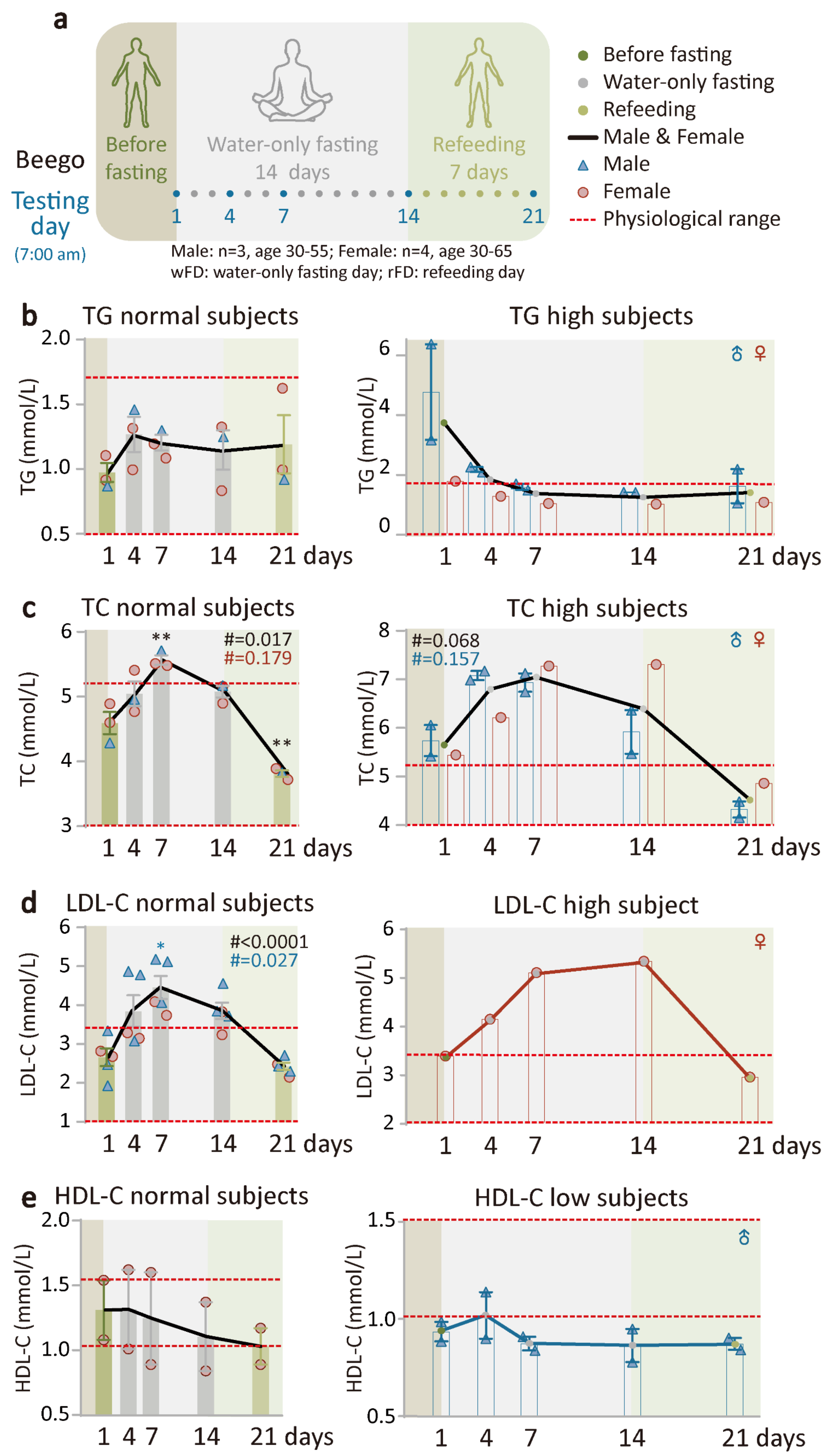


**Supplementary figure S1. Supervised beego alleviates thrombosis risk. a.** Protocol of Beego. The protocol includes fourteen water-only fasting days and seven refeeding days (rFD). Testing at 7:00-9:00 am of the following days: wFD 1, 4, 7, 14 and rFD 7 as indicated. **b**. blood triacylglyceroll (TG) test in TG normal (male=1, female=2) and high (male=2, female=1) subjects. **c**. Blood total cholesterol (TC) test in TC normal (male=1, female=2) and TC high (male=2, female=1) subjects. **d**. Low-density lipoprotein cholesterol (LDL-C) test in LDL-C normal (male=3, female=2) and high (female=1) subjects. **e**. HDL-C in the normal (female=2) and low (male=2) subjects. Data are means ± SEM. **P* < 0.05; ***P* < 0.01; ****P* < 0.001. *significant compared to the wFD1 (7:00 am) right before fasting starts .

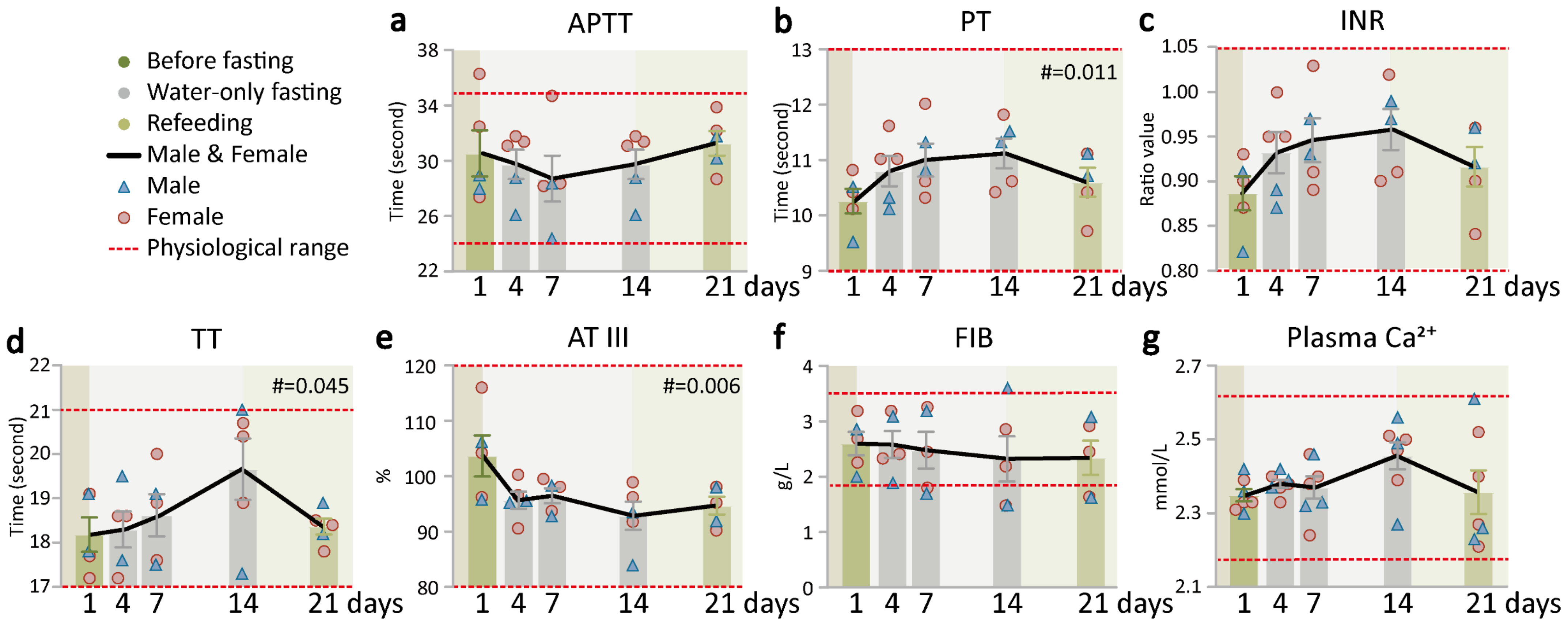


**Supplementary Figure S2 . Supervised beego maintains hemostasis capacity. a-d**. Coagulation tests. Clotting dynamics was measured at five time points (wFD1, wFD4, wFD7, wFD14 and rFD7). The tests include activated partial thromboplastin time (APTT, male=2, female=3), prothrombin time (PT, male=2, female=3), internal normalized ratio (INR, male=2, female=3), thrombin time (TT, male=2, female=3). **e-g**. Plasma levels of hemostatic biomarkers. The levels of critical proteins in hemostasis were measured at five time points as indicated. The tests include Antithrombin III (AT III, male=2, female=3); fibrinogen (FIB, male=2, female=3); $Ca^{2}$+ in plasma, male=3, female=4). Data are means ± SEM. **P* < 0.05; ***P* < 0.01; ****P* < 0.001. *significant compared to the wFD1 (7:00 am) right before fasting starts.

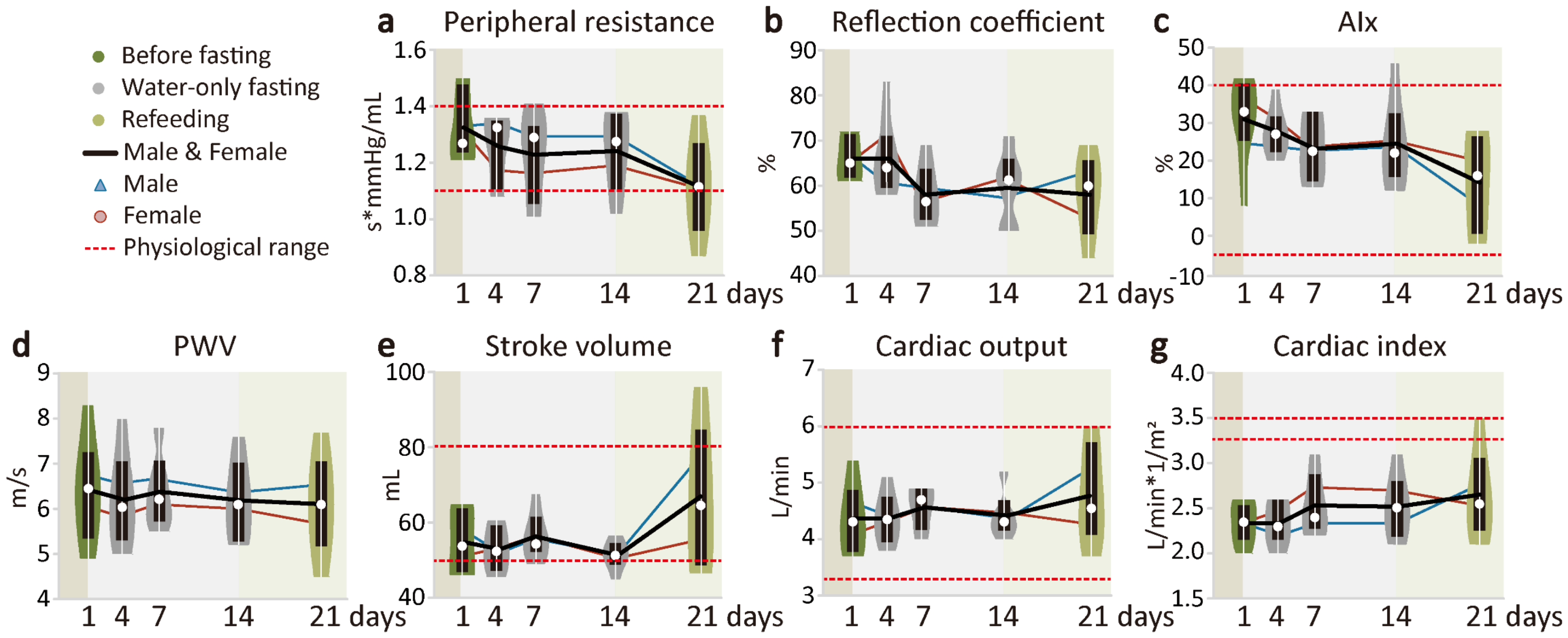


**Supplementary Figure S3. Supervised beego improves cardiovascular physiology. a-d.** Violin plots of vascular health measured with mobil-O-Graph PWA and analyzed by repeat measurement analysis and multiple comparison. Measurements include peripheral resistance (male=3, female=3), reflection coefficient (male=3, female=3), augumention index(AIx, male=3, female=3), pulse wave velocity (PWV, male=3, female=3). **e-g.** Violin plots of cardiac health measured with mobil-O-Graph PWA. Measurements include stroke volume (male=3, female=3), cardiac output (male=3, female=3) and cardiac index (male=3, female=3). Data are means ± SEM. *$P$ < 0.05; **$P$ < 0.01; ***$P$ < 0.001. *significant compared to the wFD1 (7:00 am) right before fasting starts.

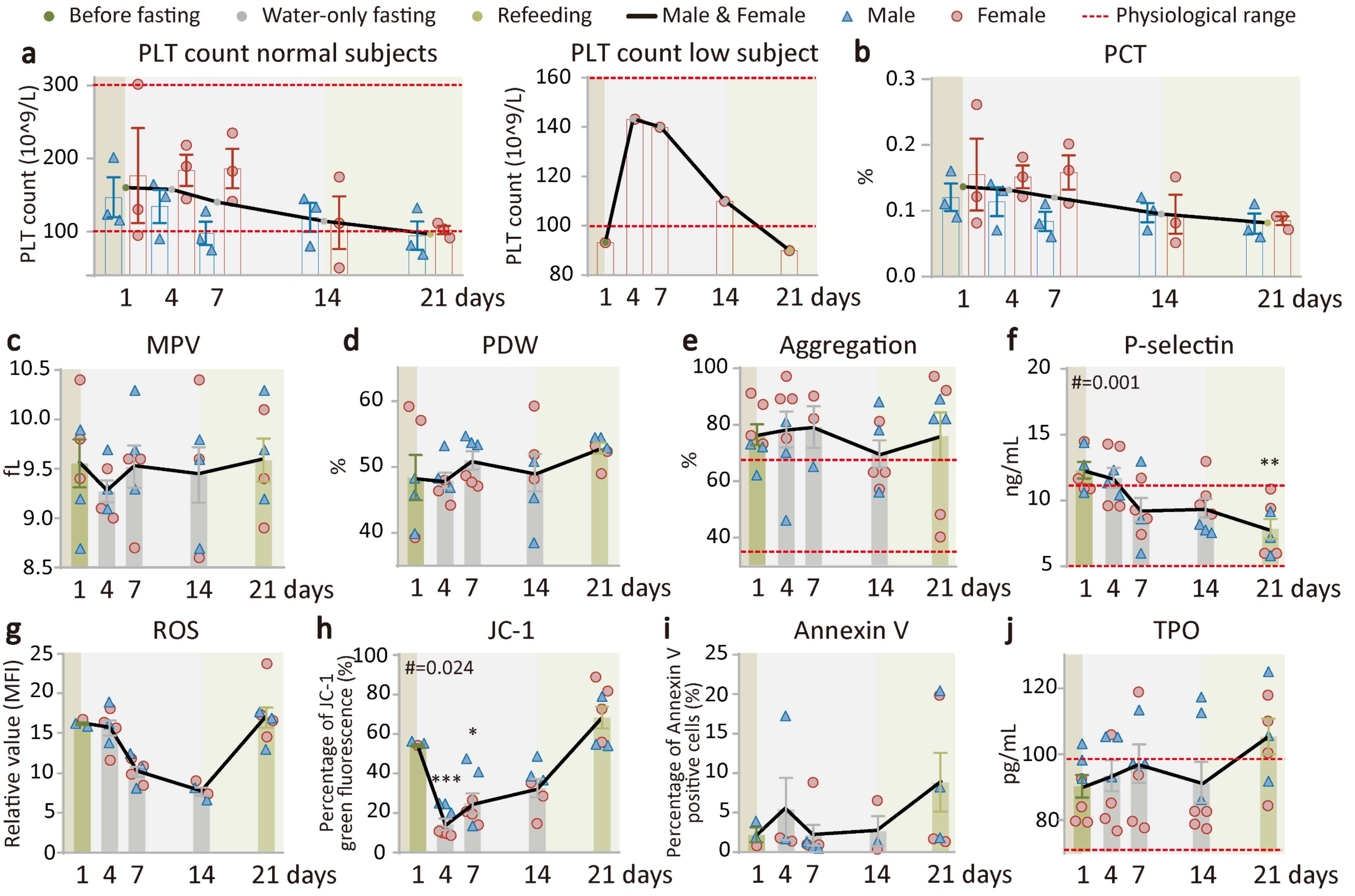


**Supplementary Figure S4. Supervised beego reduces platelet counts but retains platelet function. a**. Platelet (PLT) count in PLT normal (male=3, female=3) or low groups (female=1). **b-d**. Platelet hematocrit (PCT, male=3, female=3), Mean platelet volume (MPV, male=3, female=3), Platelet distribution width (PDW , male=3, female=3). **e.** Platelet aggregation test. The platelet aggregation after stimulation was tested at the days as indicated. male=3, female=4). **f.** ELISA measurement of P-selectin level in plasma, male=3, female=4). **g-j.** Flow cytometric measurement of platelet biology at the time points indicated**.** Total reactive oxygen species (ROS) levels in platelets (male=3, female=4); ΔΨm depolarization of mitochondria (JC-1, male=3, female=4); Percentage of Annexin V positive platelets (male=3, female=4). **j**. ELISA measurement of Thrombopoietin (TPO) level in plasma, male=3, female=4). Data are means ± SEM. *$P < 0.05$; **$P < 0.01$; ***$P < 0.001$. *significant compared to the wFD (7:00 am) right before fasting starts .

#### Refeeding Protocol

Following the water-only fasting, subjects refeed for a period of time lasting no less of half of the fasting length. We specified a specific diet and eating protocol for each of 7 refeeding days. Note that ingredient availability and typical kitchen implements are quite homogenous for most Chinese households, and counselors worked with the participants to customize serving sizes.

--Refeeding day 1 and 2:

Morning, midday, and evening: prepare a rice soup cooked with a small amount of millet by boiling the rice thoroughly with water. Only the liquid soup should be consumed (i.e., do not eat rice grains, consume no more than half a bowl of this soup at a given feeding session, but feel free to consume this soup as many as 6 or 7 times on day 1 and day 2. If it is inconvenient for you at workplace or office, you may use baby rice noodles instead. The baby rice noodles should be with millet original flavor for zero stage baby use. Don't eat too much, and do it step by step.

--Refeeding day3:

You may take vegetable soup (except spinach). For preparation the soup, wash and chop the vegetables, add water and boil; do not add oil, salt or any other seasoning. Take less veg and more soup, about half a bowl each. You may also buy baby rice noodles and eat them between meals. The baby rice noodles should be with millet original flavor for zero stage baby use. Don't eat too much, and do it step by step.

--Refeeding day 4:

You may have gruel in the morning and noodles with fresh tender vegetable leaves in the noon and evening. Make sure to boil them thoroughly. A small amount of oil and salt, but not other seasonings, may be added to the noodle. Must be tiny oil and salt, and noodles must be cooked soft. Never eat too much.

--Refeeding day 5:
For breakfast, you may have gruel or boiled noodles, with small amount of oil and salt ; you may eat boiled eggs, but only eat egg yolk. For noon meal, you may eat a small amount of rice, and rice should be soft, and eat fried green vegetables with less oil and salt. it is best to drink gruel in the evening. On the fifth day of reeating, you may add an appropriate amount of fruits, such as pears, watermelons and other fruits with more water. Don't eat too much.

--Refeeding day 6:
For breakfast, you may eat rice porridge or noodles, and boiled eggs, but only eat egg yolk. For noon and evening meal, you may eat rice (should be well done cooked) and fried green vegetables and fried tofu with less oil and salt. It is necessary to control the amount of food you take, chew food thoroughly and swallow slowly when eating. Do not drink tea and alcohol in the refeeding period.

--Refeeding day 7:
You may eat almost normally, but do not eat fried, spicy and meat food, step by step, so that your intestines and stomach can gradually return to normal function.

### Supplementary Table S1

| Index | | Gender | Mean [SEM] | | | | Adjusted *P* -value vs wFD1 | | |
|---|---|---|---|---|---|---|---|---|---|
| Index | | Gender | wFD1 | wFD4 | wFD7 | rFD7 | wFD4 | wFD7 | rFD7 |
| TG (mmol/L) | | | | | | | | | |
| | Normal | | | | | | | | |
| | | M, n=6 | 1.172[0.164] | 1.538[0.169] | 1.417[0.151] | 1.333[0.070] | | | |
| | | F, n=16 | 0.959[0.074] | 1.458[0.116] | 1.352[0.122] | 1.133[0.070] | *P* =0.006 | *P* =0.027 | *P* =0.461 |
| | High | | | | | | | | |
| | | M, n=4 | 2.788[0.535] | 2.003[0.150] | 1.713[0.142] | 1.280[0.245] | *P* =0.6730 | *P* =0.6585 | *P* =0.6585 |
| | | F, n=3 | 2.027[0.230] | 1.323[0.139] | 1.283[0.089] | 1.363[0.055] | *P* =0.029 | *P* =0.029 | *P* =0.029 |
| TC (mmol/L) | | | | | | | | | |
| | Normal | | | | | | | | |
| | | M, n=5 | 4.408[0.243] | 5.454[0.364] | 6.046[0.528] | 4.266[0.126] | *P* =0.321 | *P* =0.321 | *P* =0.993 |
| | | F, n=12 | 4.201[0.117] | 5.062[0.165] | 5.608[0.217] | 3.918[0.151] | *P* =0.003 | *P* <0.0001 | *P* =0.492 |
| | High | | | | | | | | |
| | | M, n=5 | 6.476[0.332] | 7.658[0.289] | 8.580[0.528] | 6.222[0.588] | *P* =0.2925 | *P* =0.0360 | *P* =0.9590 |
| | | F, n=7 | 5.697[0.131] | 7.210[0.313] | 7.647[0.261] | 5.197[0.195] | *P* <0.0001 | *P* <0.0001 | *P* =0.325 |
| LDL-C (mmol/L) | | | | | | | | | |
| | Normal | | | | | | | | |
| | | M, n=5 | 2.752[0.147] | 3.650[0.316] | 4.510[0.470] | 2.690[0.169] | *P* =0.2835 | *P*= 0.2250 | *P* =1.0000 |
| | | F, n=13 | 2.322[0.141] | 3.002[0.175] | 3.766[0.221] | 2.325[0.195] | *P* =0.0495 | *P* <0.0001 | *P* =1.0000 |
| | High | | | | | | | | |
| | | M, n=5 | 4.244[0.367] | 5.718[0.270] | 6.946[0.525] | 4.728[0.558] | *P* =0.1215 | *P* =0.0060 | *P* =0.7850 |
| | | F, n=6 | 3.750[0.087] | 5.025[0.345] | 5.700[0.290] | 3.465[0.182] | *P* =0.006 | *P* <0.0001 | *P* =0.754 |
| HDL-C (mmol/L) | | | | | | | | | |
| | Normal | | | | | | | | |
| | | M, n=7 | 1.183[0.063] | 1.143[0.067] | 0.931[0.057] | 0.994[0.077] | *P* =0.9500 | *P* =0.1020 | *P* =0.2025 |
| | | F, n=10 | 1.278[0.050] | 1.387[0.056] | 1.240[0.045] | 1.083[0.055] | *P* =0.4905 | *P* =0.9170 | *P* =0.0930 |
| | Low | | | | | | | | |
| | | M, n=4 | 0.938[0.033] | 0.973[0.048] | 0.793[0.067] | 0.838[0.060] | | | |
| | | F, n=1 | 1.010[0.000] | 1.550[0.000] | 1.450[0.000] | 1.030[0.000] | | | |

| | | | | | | | | |
|---|---|---|---|---|---|---|---|---|
| APTT (second) | | | | | | | | |
| | M, n=11 | 31.555[0.644] | 30.700[1.090] | 29.027[0.806] | 30.382[1.036] | | | |
| | F, n=17 | 30.400[0.719] | 29.129[0.757] | 28.124[0.770] | 29.182[0.751] | *P* =0.529 | *P* =0.282 | *P* =0.529 |
| PT (second) | | | | | | | | |
| | M, n=11 | 10.500[0.148] | 10.670[0.159] | 11.327[0.175] | 10.691[0.212] | *P* =0.841 | *P* =0.015 | *P* =0.841 |
| | F, n=17 | 10.319[0.147] | 10.876[0.176] | 11.253[0.160] | 10.312[0.126] | *P* =0.0525 | P<0.0001 | *P* =1.0000 |
| INR (ratio value) | | | | | | | | |
| | M, n=11 | 0.905[0.013] | 0.922[0.013] | 0.977[0.015] | 0.923[0.018] | *P* =0.787 | *P* =0.012 | *P* =0.787 |
| | F, n=17 | 0.889[0.013] | 0.938[0.015] | 0.970[0.014] | 0.889[0.011] | *P* =0.0495 | P<0.0001 | *P* =1.0000 |
| TT (second) | | | | | | | | |
| | M, n=11 | 18.300[0.350] | 17.660[0.246] | 17.918[0.233] | 18.382[0.308] | | | |
| | F, n=17 | 17.963[0.168] | 17.676[0.098] | 18.065[0.107] | 18.324[0.243] | | | |
| AT Ⅲ (%) | | | | | | | | |
| | M, n=11 | 97.982[2.319] | 99.900[1.615] | 97.527[1.493] | 100.891[0.906] | | | |
| | F, n=17 | 103.375[2.000] | 97.965[1.551] | 94.835[0.780] | 96.953[1.137] | *P* =0.0260 | P<0.0001 | *P* =0.0105 |
| FIB (g/L) | | | | | | | | |
| | M, n=11 | 2.505[0.265] | 3.114[0.275] | 2.895[0.158] | 2.454[0.128] | *P* =0.3960 | *P* =0.6405 | *P* =0.9960 |
| | F, n=17 | 2.458[0.099] | 2.968[0.143] | 2.645[0.121] | 2.252[0.104] | *P* =0.03 | *P* =0.55 | *P* =0.55 |
| $Ca^{2+}$ (mmol/L) | | | | | | | | |
| | M, n=11 | 2.364[0.028] | 2.429[0.025] | 2.446[0.021] | 2.337[0.042] | *P* =0.450 | *P* =0.435 | *P* =0.870 |
| | F, n=20 | 2.320[0.019] | 2.360[0.021] | 2.384[0.026] | 2.341[0.035] | | | |
| Peripheral resistance (s*mmHg/mL) | | | | | | | | |
| | M, n=10 | 1.323[0.059] | 1.248[0.032] | 1.241[0.038] | 1.241[0.070] | | | |
| | F, n=18 | 1.219[0.051] | 1.237[0.038] | 1.310[0.026] | 1.099[0.060] | *P* =1.0000 | *P* =0.8445 | *P* =0.8445 |
| Reflection coefficient (%) | | | | | | | | |
| | M, n=10 | 66.200[2.043] | 65.400[1.240] | 61.200[2.313] | 56.444[1.582] | *P* =0.9800 | *P* =0.2265 | *P* =0.0060 |
| | F, n=18 | 63.333[1.499] | 63.500[1.781] | 60.833[2.054] | 61.294[2.480] | | | |
| Alx (%) | | | | | | | | |
| | M, n=10 | 21.300[3.134] | 25.400[2.247] | 25.400[1.996] | 11.889[3.545] | *P* =0.588 | *P* =0.588 | *P* =0.180 |
| | F, n=18 | 21.222[2.285] | 29.667[2.623] | 31.667[2.752] | 17.059[2.833] | *P* =0.099 | *P* =0.051 | *P* =0.550 |
| PWV (m/s) | | | | | | | | |
| | M, n=10 | 6.320[0.318] | 6.310[0.360] | 6.070[0.361] | 6.244[0.364] | | | |

| | | | | | | | | | | |
|---|---|---|---|---|---|---|---|---|---|---|
| | | F, n=18 | | 5.883[0.248] | 5.683[0.242] | 5.872[0.242] | 5.753[0.264] | *P* =1 | *P* =1 | *P* =1 |
| Cardiac output (L/min) | | | | | | | | | | |
| | | M, n=10 | | 4.490[0.180] | 4.640[0.117] | 4.530[0.133] | 4.633[0.172] | | | |
| | | F, n=18 | | 4.394[0.213] | 4.028[0.108] | 4.106[0.113] | 4.706[0.230] | *P* =0.897 | *P* =0.897 | *P* =0.897 |
| Cardiac index (L/min*1/m$^2$) | | | | | | | | | | |
| | | M, n=10 | | 2.420[0.104] | 2.510[0.080] | 2.460[0.095] | 2.500[0.118] | | | |
| | | F, n=18 | | 2.667[0.113] | 2.444[0.081] | 2.572[0.077] | 2.876[0.132] | *P* =0.519 | *P* =0.850 | *P* =0.519 |
| Blood pressure (mmHg) | | | | | | | | | | |
| | Left | | | | | | | | | |
| | | F, n=1 | | | | | | | | |
| | | | Sbp | 143 | 114 | 124 | 119 | | | |
| | | | Dbp | 107 | 77 | 87 | 81 | | | |
| | Middle | | | | | | | | | |
| | | M, n=1 | | | | | | | | |
| | | | Sbp | 141 | 147 | 144 | 136 | | | |
| | | | Dbp | 94 | 94 | 93 | 89 | | | |
| | | F, n=2 | | | | | | | | |
| | | | Sbp | 156[6.000] | 137[9.500] | 146[3.000] | 131[6.000] | | | |
| | | | Dbp | 89[1.500] | 79[0.000] | 87[5.000] | 77.5[3.500] | | | |
| | Right | | | | | | | | | |
| | | M, n=10 | | | | | | | | |
| | | | Sbp | 125.1[3.104] | 122.9[3.086] | 121.6[2.845] | 122.3[3.347] | | | |
| | | | Dbp | 74.2[2.607] | 73.4[2.701] | 73.9[2.132] | 73.3[2.642] | | | |
| | | F, n=18 | | | | | | | | |
| | | | Sbp | 112.611[3.192] | 108.722[2.799] | 113.944[3.234] | 110.556[2.862] | | | |
| | | | Dbp | 67.944[3.110] | 64.000[2.265] | 72.333[2.036] | 65.111[2.475] | *P* =0.887 | *P* =0.887 | *P* =0.887 |
| PLT (10^9/L) | | | | | | | | | | |
| | Normal | | | | | | | | | |
| | | M, n=11 | | 154.000[10.015] | 106.364[8.883] | 128.091[15.246] | 94.818[11.144] | *P* =0.024 | *P* =0.278 | *P* =0.006 |
| | | F, n=20 | | 142.900[13.913] | 132.500[10.956] | 121.550[10.196] | 93.632[9.327] | *P* =0.8510 | *P* =0.5985 | *P* =0.0240 |

| | | | | | | | | |
|---|---|---|---|---|---|---|---|---|
| | Low | | | | | | | |
| | F, n=5 | 61.111[11.205] | 122.444[29.375] | 109.333[27.872] | 92.222[25.196] | | | |
| PCT (%) | | | | | | | | |
| | M, n=11 | 0.146[0.009] | 0.103[0.007] | 0.122[0.014] | 0.090[0.010] | *P* =0.0255 | *P* =0.2610 | *P* =0.0060 |
| | F, n=20 | 0.135[0.012] | 0.128[0.010] | 0.116[0.009] | 0.093[0.008] | *P* =0.9250 | *P* =0.6195 | *P* =0.0360 |
| MPV (fL) | | | | | | | | |
| | M, n=11 | 9.355[0.142] | 9.782[0.165] | 9.455[0.122] | 9.745[0.167] | | | |
| | F, n=20 | 9.560[0.192] | 9.750[0.151] | 9.600[0.167] | 10.026[0.211] | *P* =0.997 | *P* =0.997 | *P* =0.540 |
| PDW (%) | | | | | | | | |
| | M, n=11 | 45.955[1.017] | 50.891[1.529] | 49.809[1.753] | 52.236[1.030] | *P* =0.0585 | *P* =0.1320 | *P* =0.0210 |
| | F, n=20 | 45.770[1.669] | 48.325[1.074] | 50.735[1.221] | 51.337[1.505] | *P* =0.419 | *P* =0.051 | *P* =0.051 |
| Aggregation (%) | | | | | | | | |
| | M, n=11 | 70.600[4.966] | 64.444[6.594] | 81.800[5.825] | 67.143[7.732] | | | |
| | F, n=20 | 74.200[9.332] | 75.533[5.012] | 87.722[2.843] | 70.154[5.938] | | | |
| P-selectin (ng/mL) | | | | | | | | |
| | M, n=11 | 12.341[0.634] | 10.454[0.678] | 9.199[0.558] | 9.185[0.538] | *P* =0.084 | *P* =0.003 | *P* =0.003 |
| | F, n=19 | 11.359[0.418] | 10.699[0.458] | 9.749[0.430] | 9.574[0.479] | *P* =0.5950 | *P* =0.0525 | *P* =0.0510 |
| ROS (MFI) | | | | | | | | |
| | M, n=5 | 18.290[1.466] | 12.236[0.952] | 10.034[1.311] | 8.518[0.668] | *P* =0.0070 | *P* =0.0015 | *P* <0.0001 |
| | F, n=5 | 17.092[0.658] | 14.894[0.962] | 7.378[1.087] | 8.684[0.327] | *P* =0.175 | *P* <0.0001 | *P* <0.0001 |
| JC-1 (%) | | | | | | | | |
| | M, n=5 | 57.097[1.911] | 17.996[3.308] | 21.794[3.116] | 28.378[5.076] | *P* <0.0001 | *P* <0.0001 | *P* =0.001 |
| | F, n=5 | 57.148[2.009] | 41.102[10.411] | 44.160[14.892] | 33.844[4.403] | | | |
| Annexin V (%) | | | | | | | | |
| | M, n=5 | 6.886[3.372] | 3.280[2.089] | 2.822[1.179] | 0.960[0.670] | | | |
| | F, n=5 | 1.286[0.260] | 3.118[1.232] | 3.896[2.226] | 0.720[0.163] | | | |
| TPO (pg/mL) | | | | | | | | |
| | M, n=11 | 94.155[5.821] | 97.658[3.387] | 101.340[5.207] | 95.152[3.613] | | | |
| | F, n=20 | 92.113[3.408] | 94.344[3.470] | 98.447[3.501] | 98.765[3.479] | | | |

#### Supplementary Table S2

| Index | | Gender | Mean [SEM] | | | | | Adjusted *P* -value vs wFD1 | | | |
|---|---|---|---|---|---|---|---|---|---|---|---|
| | | | wFD1 | wFD4 | wFD7 | wFD14 | rFD7 | wFD4 | wFD7 | wFD14 | rFD7 |
| TG (mmol/L) | | | | | | | | | | | |
| | Normal | | | | | | | | | | |
| | | M, n=1 | 0.870 | 1.460 | 1.300 | 1.250 | 0.920 | | | | |
| | | F, n=2 | 1.015[0.095] | 1.160[0.160] | 1.145[0.055] | 1.085[0.245] | 1.315[0.315] | | | | |
| | High | | | | | | | | | | |
| | | M, n=2 | 4.740[1.600] | 2.145[0.085] | 1.565[0.105] | 1.385[0.005] | 1.590[0.570] | | | | |
| | | F, n=1 | 1.750 | 1.250 | 1.010 | 0.990 | 1.050 | | | | |
| TC (mmol/L) | | | | | | | | | | | |
| | Normal | | | | | | | | | | |
| | | M, n=1 | 4.270 | 4.940 | 5.690 | 5.150 | 3.810 | | | | |
| | | F, n=2 | 4.725[0.145] | 5.070[0.320] | 5.475[0.015] | 5.010[0.130] | 3.785[0.085] | | | | |
| | High | | | | | | | | | | |
| | | M, n=2 | 5.720[0.320] | 7.045[0.095] | 6.900[0.190] | 5.900[0.450] | 4.305[0.165] | | | | |
| | | F, n=1 | 5.410 | 6.200 | 7.240 | 7.290 | 4.850 | | | | |
| LDL-C (mmol/L) | | | | | | | | | | | |
| | Normal | | | | | | | | | | |
| | | M, n=3 | 2.600[0.411] | 4.263[0.582] | 4.810[0.360] | 4.063[0.261] | 2.500[0.121] | *P* =0.0720000[illegible] | *P* = 0.0280000[illegible] | *P* =0.0866666[illegible] | *P* =0.99900000 |
| | | F, n=2 | 2.770[0.070] | 3.240[0.070] | 3.935[0.175] | 3.555[0.295] | 2.335[0.165] | | | | |
| | High | | | | | | | | | | |
| | | F, n=1 | 3.37 | 4.13 | 5.09 | 5.32 | 2.94 | | | | |
| HDL-C (mmol/L) | | | | | | | | | | | |
| | Normal | | | | | | | | | | |
| | | F, n=2 | 1.310[0.230] | 1.315[0.305] | 1.245[0.355] | 1.105[0.265] | 1.030[0.140] | | | | |
| | Low | | | | | | | | | | |
| | | M, n=2 | 0.940[0.050] | 1.020[0.120] | 0.875[0.035] | 0.865[0.085] | 0.870[0.030] | | | | |
| APTT (second) | | | | | | | | | | | |
| | | M, n=2 | 28.400[0.500] | 27.350[1.350] | 26.300[2.000] | 27.350[1.350] | 30.900[0.800] | | | | |
| | | F, n=3 | 31.967[2.578] | 31.333[0.203] | 30.333[2.134] | 31.333[0.203] | 31.500[1.531] | | | | |
| PT (second) | | | | | | | | | | | |
| | | M, n=2 | 10.000[0.500] | 10.200[0.100] | 11.050[0.250] | 11.400[0.100] | 10.900[0.200] | | | | |
| | | F, n=3 | 10.433[0.203] | 11.200[0.200] | 10.967[0.524] | 10.933[0.437] | 10.400[0.404] | | | | |
| INR (ratio value) | | | | | | | | | | | |
| | | M, n=2 | 0.865[0.045] | 0.880[0.010] | 0.950[0.020] | 0.980[0.010] | 0.940[0.020] | | | | |
| | | F, n=3 | 0.900[0.017] | 0.967[0.017] | 0.943[0.044] | 0.943[0.038] | 0.900[0.035] | | | | |
| TT (second) | | | | | | | | | | | |
| | | M, n=2 | 18.450[0.650] | 18.550[0.950] | 18.300[0.800] | 19.150[1.850] | 18.550[0.350] | | | | |

| | | | | | | |
|---|---|---|---|---|---|---|
| | F, n=3 | 18.000[0.569] | 18.133[0.467] | 18.833[0.694] | 20.000[0.557] | 18.233[0.219] |
| AT Ⅲ (%) | | | | | | |
| | M, n=2 | 101.000[5.200] | 95.400[0.200] | 95.550[2.750] | 88.700[4.800] | 94.950[3.050] |
| | F, n=3 | 105.467[5.751] | 95.833[2.826] | 97.100[1.747] | 95.633[2.069] | 94.500[2.307] |
| FIB (g/L) | | | | | | |
| | M, n=2 | 2.430[0.430] | 2.490[0.600] | 2.445[0.745] | 2.545[1.055] | 2.350[0.730] |
| | F, n=3 | 2.713 [0.269] | 2.643[0.274] | 2.503[0.422] | 2.177[0.398] | 2.337[0.374] |
| $Ca^{2+}$ (mmol/L) | | | | | | |
| | M, n=3 | 2.360[0.035] | 2.393[0.015] | 2.370[0.045] | 2.440[0.087] | 2.367[0.122] |
| | F, n=4 | 2.340 [0.017] | 2.370[0.015] | 2.370[0.047] | 2.468[0.027] | 2.350[0.069] |
| Peripheral resistance (s*mmHg/mL) | | | | | | |
| | M, n=3 | 1.330[0.071] | 1.340[0.012] | 1.293[0.007] | 1.293[0.082] | 1.120[0.144] |
| | F, n=3 | 1.320[0.091] | 1.173[0.079] | 1.163[0.125] | 1.190[0.087] | 1.110[0.069] |
| Reflection coefficient (%) | | | | | | |
| | M, n=3 | 66.667[2.603] | 60.667[1.764] | 59.667[5.207] | 57.333[3.712] | 63.000[3.000] |
| | F, n=3 | 65.333[3.383] | 71.333[5.897] | 56.333[2.848] | 61.667[6.173] | 53.000[5.859] |
| Alx (%) | | | | | | |
| | M, n=3 | 24.667[8.413] | 23.667[2.333] | 22.667[5.364] | 23.667[3.383] | 8.333[8.876] |
| | F, n=3 | 37.333[3.283] | 31.333[3.930] | 23.667[5.812] | 25.333[10.477] | 20.000[4.619] |
| PWV (m/s) | | | | | | |
| | M, n=3 | 6.767[0.819] | 6.567[0.762] | 6.667[0.593] | 6.367[0.669] | 6.533[0.664] |
| | F, n=3 | 6.067[0.601] | 5.833[0.491] | 6.100[0.379] | 6.000[0.462] | 5.667[0.664] |
| Stroke volume (mL) | | | | | | |
| | M, n=3 | 58.050[6.007] | 52.333[3.784] | 55.980[1.766] | 51.853[1.050] | 78.123[11.174] |
| | F, n=3 | 51.477[4.257] | 53.557[3.789] | 56.687[5.594] | 50.810[3.366] | 55.790[7.899] |
| Cardiac output (L/min) | | | | | | |
| | M, n=3 | 4.633[0.463] | 4.400[0.379] | 4.567[0.203] | 4.367[0.088] | 5.267[0.546] |
| | F, n=3 | 4.100[0.231] | 4.333[0.176] | 4.567[0.285] | 4.467[0.371] | 4.267[0.318] |
| Cardiac index (L/min*1/$m^2$) | | | | | | |
| | M, n=3 | 2.333[0.133] | 2.200[0.115] | 2.333[0.067] | 2.333[0.186] | 2.767[0.406] |
| | F, n=3 | 2.333[0.167] | 2.467[0.133] | 2.733[0.233] | 2.700[0.200] | 2.533[0.186] |

| | | | | | | | |
|---|---|---|---|---|---|---|---|
| PLT (10^9/L) | | | | | | | |
| | Normal | | | | | | |
| | | M, n=3 | 145.556[27.315 | 32.963[22.556 | 96.296[15.835] | 118.148[19.890 | 92.963[19.398] |
| | | F, n=3 | 175.185[64.891 | 82.593[21.328 | 184.815[27.007] | 110.741[35.926 | 100.741[5.821] |
| | Low | | | | | | |
| | | F, n=1 | 93.333 | 143.333 | 140.000 | 110.000 | 90.000 |
| PCT (%) | | | | | | | |
| | | M, n=3 | 0.120[0.021] | 0.113[0.022] | 0.083[0.015] | 0.097[0.015] | 0.080[0.015] |
| | | F, n=3 | 0.153[0.055] | 0.150[0.017] | 0.157[0.026] | 0.093[0.030] | 0.083[0.007] |
| MPV (fL) | | | | | | | |
| | | M, n=3 | 9.267[0.348] | 9.367[0.176] | 9.767[0.291] | 9.367[0.338] | 9.733[0.318] |
| | | F, n=3 | 9.867[0.291] | 9.200[0.153] | 9.300[0.300] | 9.533[0.521] | 9.467[0.348] |
| PDW (%) | | | | | | | |
| | | M, n=3 | 44.567[2.490] | 49.367[1.910] | 53.833[0.393] | 44.767[3.557] | 53.867[0.533] |
| | | F, n=3 | 51.867[6.313] | 46.200[1.102] | 47.833[0.467] | 53.133[3.263] | 51.533[1.288] |
| Aggregation (%) | | | | | | | |
| | | M, n=3 | 69.000[3.512] | 65.667[10.333] | 65.000[0.000] | 74.000[9.452] | 84.333[2.333] |
| | | F, n=4 | 81.750[4.308] | 87.500[4.573] | 86.000[4.000] | 66.000[5.196] | 69.250[14.705] |
| P-selectin (ng/mL) | | | | | | | |
| | | M, n=3 | 12.567[1.093] | 11.353[0.551] | 9.183[2.042] | 7.833[0.188] | 7.380[0.966] |
| | | F, n=4 | 11.963[0.851] | 11.865[1.334] | 9.230[0.873] | 10.463[0.873] | 8.038[1.236] |
| ROS (MFI) | | | | | | | |
| | | M, n=3 | 16.315[0.175] | 16.537[1.502] | 10.600[1.265] | 7.560[0.770] | 16.100[1.473] |
| | | F, n=4 | 16.730[0.000] | 15.428[1.382] | 10.198[0.728] | 8.135[0.825] | 18.073[1.999] |
| JC-1 (%) | | | | | | | |
| | | M, n=3 | 54.145[0.495] | 21.553[1.575] | 32.240[10.423] | 39.673[3.753] | 60.943[8.180] |
| | | F, n=4 | 52.460[0.000] | 7.960[0.447] | 18.430[2.585] | 24.437[6.111] | 73.078[7.130] |
| Annexin V (%) | | | | | | | |
| | | M, n=3 | 3.040[0.990] | 9.580[7.790] | 0.923[0.245] | 1.450 [0.000] | 10.283[5.460] |
| | | F, n=4 | 0.770[0.000] | 1.550[0.200] | 3.547[2.647] | 3.415[3.115] | 7.627[6.172] |
| TPO (pg/mL) | | | | | | | |
| | | M, n=3 | 97.817[3.070] | 101.037[4.068] | 102.387[5.432] | 105.203 [9.685] | 107.360[9.660] |
| | | F, n=4 | 84.055[3.077] | 87.220[6.519] | 92.620[9.515] | 80.520[1.359] | 103.363[7.226] |
